# A 5-transcript signature for discriminating viral and bacterial etiology in pediatric pneumonia

**DOI:** 10.1101/2024.11.04.24316062

**Authors:** Sandra Viz-Lasheras, Alberto Gómez-Carballa, Jacobo Pardo-Seco, Xabier Bello, Irene Rivero-Calle, Ana Isabel Dacosta, Myrsini Kaforou, Dominic Coote, Aubrey J. Cunnington, Marieke Emonts, Jethro A. Herberg, Victoria J. Wright, Enitan D. Carrol, Stephane C. Paulus, Werner Zenz, Daniela S. Kohlfürst, Michiel Van der Flier, Ronald de Groot, Luregn J. Schlapbach, Philipp Agyeman, Andrew J. Pollard, Colin Fink, Taco T. Kuijpers, Suzanne Anderson, Cristina Calvo, María del Carmen Martínez-Padilla, Ana Pérez-Aragón, Esteban Gómez-Sánchez, Juan Valencia-Ramos, Francisco Giménez-Sánchez, Paula Alonso-Quintela, Laura Moreno-Galarraga, Ulrich von Both, Marko Pokorn, Dace Zavadska, María Tsolia, Clementien L. Vermont, Henriëtte A. Moll, Michael Levin, Federico Martinón-Torres, Antonio Salas EUCLIDS, DIAMONDS, GENDRES, and PERFORM consortia

## Abstract

Pneumonia stands as the primary cause of death among children under five, yet current diagnosis methods often result in inadequate or unnecessary treatments. Our research seeks to address this gap by identifying host transcriptomic biomarkers in the blood of children with definitive viral and bacterial pneumonia. We performed RNA sequencing on 192 prospectively collected whole blood samples, including 38 controls and 154 pneumonia cases, uncovering a 5-transcript signature (genes *FAM20A*, *BAG3*, *TDRD9*, *MXRA7* and *KLF14*) that effectively distinguishes bacterial from viral pneumonia (AUC: 0.95 [0.88–1.00]) Initial validation using combined definitive and probable cases yielded an AUC of 0.87 [0.77–0.97], while full validation in a new prospective cohort of 32 patients achieved an AUC of 0.92 [0.83–1]. This robust signature holds significant potential to enhance diagnostics accuracy for pediatric pneumonia, reducing diagnostic delays and unnecessary treatments, and potentially transforming clinical practice.

## Introduction

Pneumonia is an acute respiratory infection, and one of the leading causes of morbidity and mortality worldwide. In children younger than 5 (excluding the neonatal period), it remains the primary cause of death ^1,2^. Common symptoms in children include fever, tachypnoea,cough, chest pain and difficulty in breathing ^3^. Pneumonia can be caused by a variety of microorganisms, including bacteria, viruses and fungi, depending on age groups and the specific epidemiological situation. However, *Streptococcus pneumoniae* is the most common bacterial cause of pneumonia across all age group ^2,4^. The causative microorganisms also vary significantly between the two main types of pneumonia: Community-Acquired Pneumonia (CAP) and Hospital-Acquired Pneumonia (HAP). *Pseudomonas aeruginosa*, *Staphylococcus aureus*, and *Enterobacter* are the most common causes of HAP ^5^ whereas *Streptococcus pneumoniae*, *Staphylococcus aureus*, *Streptococcus pyogenes*, *Haemophilus influenzae*, *Mycoplasma pneumoniae*, *Legionella pneumophila* and respiratory viruses, such as *Respiratory Syncytial virus*, *Rhinovirus* or *Influenza virus A/B*, are the most common microorganisms responsible for CAP ^3,6^. The introduction of improved conjugate vaccines against *Haemophilus influenzae* type b and *Streptococcus pneumoniae* has led to a reduction in the incidence and severity of childhood pneumonia, resulting in significant changes in the proportions of CAP cases etiologies ^7,8^.

The clinical presentation of pneumonia is diverse and may overlap with other respiratory conditions such as asthma, bronchiolitis, pertussis, lung abscess, bronchiectasis, or malaria, among others ^6,9–14^. Accurate assessment of disease severity ^15^ is crucial for effective management, influencing decisions on antibiotic prescriptions and hospitalization ^16,17^. Despite efforts to enhance international guidelines ^9,10,18^, the variability in causative microorganisms and symptoms poses a significant challenge for clinicians in healthcare centers ^14,19–22^. Current diagnostic criteria rely on nonspecific symptoms, chest imaging, and inconclusive laboratory analysis ^14,19,23,24^, with limitations in differentiating the etiology of the disease (i.e., viral, bacterial). Obtaining lower respiratory tract samples in children is often difficult ^19,20^. Consequently, misdiagnosis of bacterial pneumonia is common, leading to increased costs, unnecessary medical tests, hospital admissions, and incorrect antibiotic prescriptions ^6,25^.

Giving these challenges, numerous studies have suggested the potential use of biomarkers as supplementary tools in the managing and diagnosis of pneumonia ^9,16,25–27^. Serum or plasma C-reactive protein (CRP) and procalcitonin (PCT) have been identified as potential biomarkers to assist in the diagnosis and prognosis of pneumonia patients ^9,16,28,29^.. The study of host transcriptomic in the field of infectious disease has become increasingly important in recent years, leading to advancements in understanding host-pathogen interactions and the development of useful tools for diagnosing and prognosing diseases, including the development of point-of care devices ^30–35^. There are a growing number of studies investigating host gene expression biomarkers related to different infectious diseases, such as tuberculosis ^36–38^, H1N1 ^39^, RSV ^40^, rotavirus ^41^ and also aiming at differentiating bacterial from viral infection ^30,35,42,43^. However, the study of host transcriptomics in pneumonia patients has not received as much attention, and only a few studies describing blood transcriptomic biomarkers related to CAP in adults are available in the literature ^27,44^. Some studies have recently focused on pneumonia outcome prediction ^45–47^ or pneumonia etiology discrimination ^48,49^.

Regarding pediatric pneumonia, most studies have focused on the potential use of already described biomarkers, such as white blood cell (WBC) count, neutrophil percentage (NP), serum CRP and PCT. However, results indicate that these biomarkers are not good predictors for pediatric pneumonia due to their inability to differentiate pneumonia from bacterial and viral origins ^50,51^. To our knowledge, there is only one study investigating blood transcriptomics in the context of pediatric pneumonia etiology, but in a very specific population from a malaria-endemic area and using a signature with a high number of genes ^52^. Therefore, further studies are necessary to approach this complex disease in children.

In light of these challenges in pneumonia diagnosis and management in children, we analyze the whole blood transcriptome of the largest pediatric pneumonia cohort recruited to date with the aim of *i*) shedding light on the molecular mechanisms underlying pediatric pneumonia, providing insights of new therapeutic targets; *ii*) investigating host’s differential response to pneumonia of bacterial and viral origin; and *iii*) proposing a minimal transcriptomic signature that allows for the differentiation between viral and bacterial pneumonia in children.

## Results

### Discovery cohort description

In this study, blood samples were collected from 192 children (see **Figure 1** and methods for details on the experimental design). Among them, 154 were hospitalized with pneumonia (median age: 3.4 years; 51.3% male) and 38 were healthy controls (median age: 3.3 years; 68.4% male). Clinical and demographic details of the entire cohort and sub-categories used in different analyses are provided in **Table 1**. Among the pneumonia patients, 25% were admitted to PICU, 3% died, and 39% required oxygen support (16.2% with invasive ventilation). No statistically significant differences were found between the bacterial and viral pneumonia sub-groups for these parameters. The majority of bacterial pneumonias were caused by *Streptococcus pneumoniae* (47.5%) followed by *Staphylococcus aureus* (10.0%*) and Streptococcus pyogenes* (7.5%) *between others* (35.0%). Viral pneumonias were caused by influenza virus (22.2%), bocavirus (22.2%), respiratory syncytial virus (22.2%), rhinovirus (11.1%), adenovirus (11.1%) and parainfluenza (11.1%).

**Figure 1.**
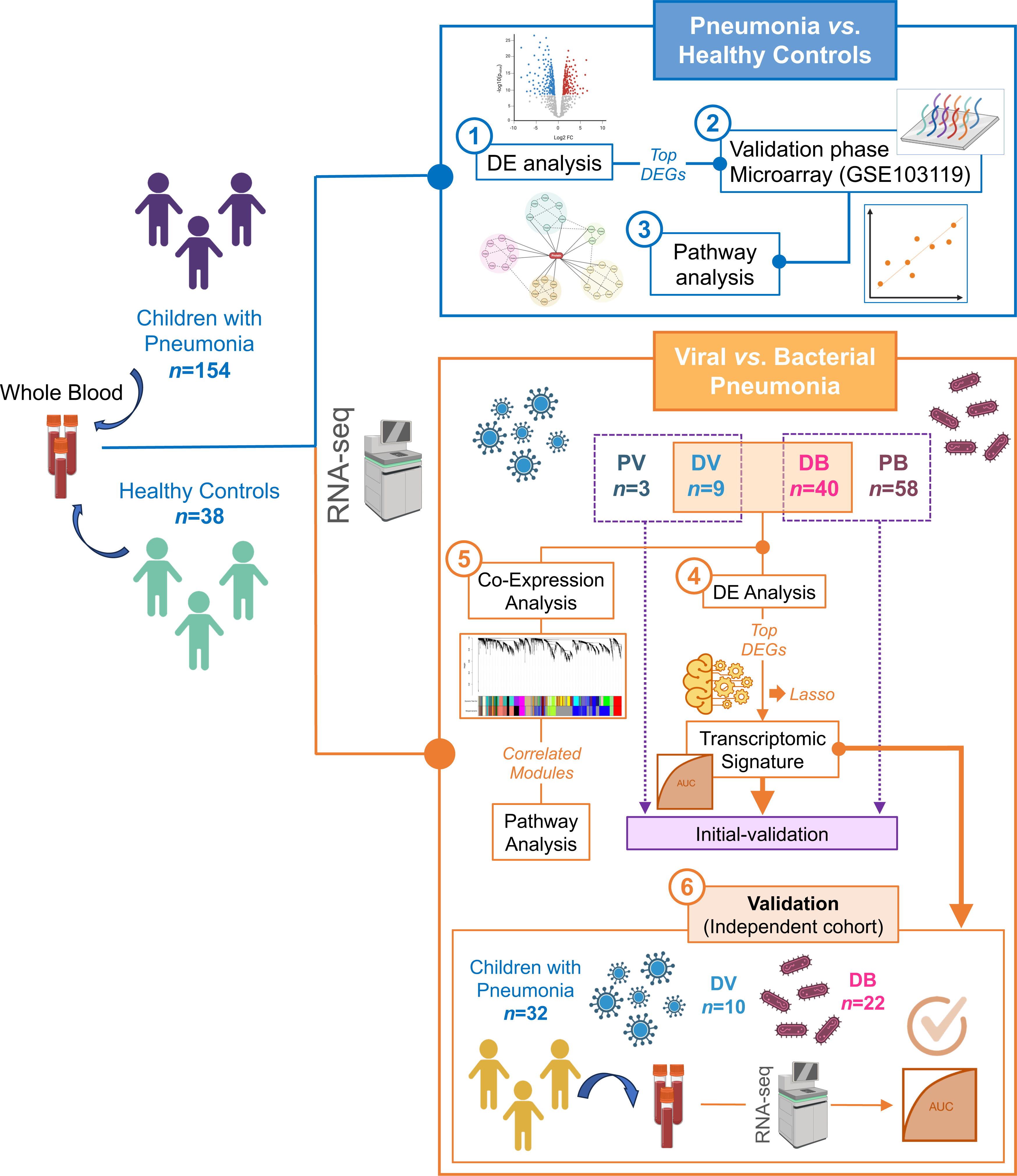
Scheme of the study design. The figure was built using Biorender resources; *Created in BioRender. lab 19, G. (2024) BioRender.com/b32x666*.

**Table 1.** Clinical features of the pneumonia patient’s cohort. Wilcoxon and Fisher exact test were used to assess statistical significance between groups in numerical and categorical variables respectively. IV: invasive ventilation; NIV: non-invasive ventilation (*) indicates statistical significance.

|  | Pneumonia vs. Healthy controls |  |  | DV vs. DB |  |  |  |  | DV + PV vs. DB + PB |  |  |  |  |
| --- | --- | --- | --- | --- | --- | --- | --- | --- | --- | --- | --- | --- | --- |
|  | Controls<br>(n = 38) | Pneumonia<br>(n = 154) | n | DB<br>(n = 40) | n | DV<br>(n = 9) | n | P-value | DB + PB<br>(n = 98) | n | DV + PV<br>(n = 12) | n | P-value |
| Gender (Male) | 68.4% (36/38) | 51.3% (79/154) | 154 | 62.5% (25/40) | 40 | 55.6% (5/9) | 9 | 0.72 | 53.1% (52/98) | 98 | 58.3% (7/12) | 12 | 0.769 |
| Age (years) | 3.3 (2.3-8.2) | 3.36 (1.24-6.17) | 153 | 2.77 (1.25-5.78) | 40 | 2.75 (1.52-4.76) | 8 | 0.923 | 3.78 (1.54-6.285) | 98 | 1.93 (0.94-3.88) | 11 | 0.096 |
| Days hospitalized | – | 5 (3-10) | 149 | 6.5 (3.5-14) | 38 | 6 (4.75-8.5) | 8 | 0.621 | 5 (3-9) | 95 | 6 (3.5-9) | 11 | 0.536 |
| PICU admission | – | 24.7% (38/154) | 154 | 30% (12/40) | 40 | 22.2% (2/9) | 9 | 1 | 22.4% (22/98) | 98 | 25% (3/12) | 12 | 1 |
| PICU days | – | 7 (3-11) | 37 | 11 (6.5-16) | 11 | 6.5 (4.25-8.75) | 2 | 0.372 | 6 (3-11) | 21 | 2 (2-6.5) | 3 | 0.356 |
| Death | – | 3.2% (5/154) | 154 | 7.5% (3/40) | 40 | 11.1% (1/9) | 9 | 0.569 | 4.1% (4/98) | 98 | 8.3% (1/12) | 12 | 0.445 |
| Treatments |  |  |  |  |  |  |  |  |  |  |  |  |  |
| Inotropes | – | 9.7% (15/154) | 154 | 5% (2/40) | 40 | 11.1% (1/9) | 9 | 0.464 | 6.1% (6/98) | 98 | 8.3% (1/12) | 12 | 0.565 |
| IV | – | 16.2% (25/154) | 154 | 15% (6/40) | 40 | 22.2% (2/9) | 9 | 0.628 | 11.2% (11/98) | 98 | 25% (3/12) | 12 | 0.18 |
| NIV | – | 11% (17/154) | 154 | 15% (6/40) | 40 | 11.1% (1/9) | 9 | 1 | 8.2% (8/98) | 98 | 8.3% (1/12) | 12 | 1 |
| Oxygen | – | 39% (60/154) | 154 | 40% (16/40) | 40 | 33.3% (3/9) | 9 | 1 | 34.7% (34/98) | 98 | 41.7% (5/12) | 12 | 0.751 |
| Analytical parameters |  |  |  |  |  |  |  |  |  |  |  |  |  |
| CRP (mg/L) | – | 81 (30.1-215.5) | 59 | 146 (34.5-244) | 11 | 28.5 (11.85-30.15) | 6 | 0.062 | 190 (81-273) | 33 | 27 (5.9-30.1) | 7 | 0.001* |
| Platelets (10<sup>9</sup>/L) | – | 322 (259-433) | 135 | 287.5 (175.5-405.5) | 28 | 491 (283-507) | 5 | 0.173 | 324 (262.75-421.5) | 84 | 398 (273.5-495) | 8 | 0.458 |
| White Cells (10<sup>9</sup>/L) | – | 17.2 (10.2-25) | 133 | 18.75 (10.075-26.075) | 28 | 7.2 (6.35-16.25) | 3 | 0.423 | 18.65 (12.18-26.72) | 84 | 6.85 (5.75-20.775) | 6 | 0.089 |
| Lactate (mmol/L) | – | 1.9 (1.18-2.65) | 30 | 2.5 (2.3-2.7) | 5 | 2.5 (2.5-2.5) | 1 | 1 | 2.2 (1.48-2.55) | 16 | 2.5 (2.5-2.5) | 1 | 0.539 |
| Neutrophils (10<sup>9</sup>/L) | – | 11.87 (6.47-19.95) | 128 | 13.4 (8.05-20.88) | 26 | 12.06 (7.94-16.18) | 2 | 0.762 | 14.61 (7.82-23.62) | 82 | 4.16 (3.82-18.6) | 5 | 0.111 |
| Monocytes (10<sup>9</sup>/L) | – | 0.8 (0.17-1.7) | 84 | 0.61 (0.125-1.2) | 14 | 0.8 (0.4-1.15) | 3 | 0.801 | 0.955 (0.168-1.92) | 56 | 0.02 (0-0.8) | 5 | 0.068 |
| Lymphocytes (10<sup>9</sup>/L) | – | 3.17 (1.67-5.4) | 130 | 1.95 (1.4-5.65) | 27 | 4.1 (3.42-8.55) | 3 | 0.213 | 2.6 (1.5-5.4) | 83 | 3.42 (2.1475-6.35) | 6 | 0.381 |

As expected, C-reactive protein (CRP) values were significantly higher in DB + PB compared to DV + PV (*P*-value = 0.001). CRP values were higher in the DB group compared to DV; this difference approached the significance threshold (*P*-value = 0.062), likely due to limited statistical power, especially given the low number of samples in DV group, coupled with missing data. For the remaining blood tests, there were no significant statistical differences between bacterial and viral sub-groups.

### Differentially expressed genes in pneumonia

Through a comparative analysis of transcriptomes in children with pneumonia *vs*. healthy controls, we identified 5,486 differentially expressed genes (DEGs), using a significance threshold of Benjamini-Hochberg False Discovery Rate (FDR) 5% (see **Table S1**). Within this set of DEGs, 2,716 were found to be upregulated, while 2,770 were downregulated.

To gain insight into the overall variation in gene expression, we performed a principal component analysis (PCA) on a subset of 100 highly variable genes out of a total of 192 samples. The first principal component (PC1), explaining approximately 30% of the variation, effectively segregated the samples into two main clusters, corresponding to pneumonia samples and healthy control samples (**Figure 2A**).

**Figure 2.**
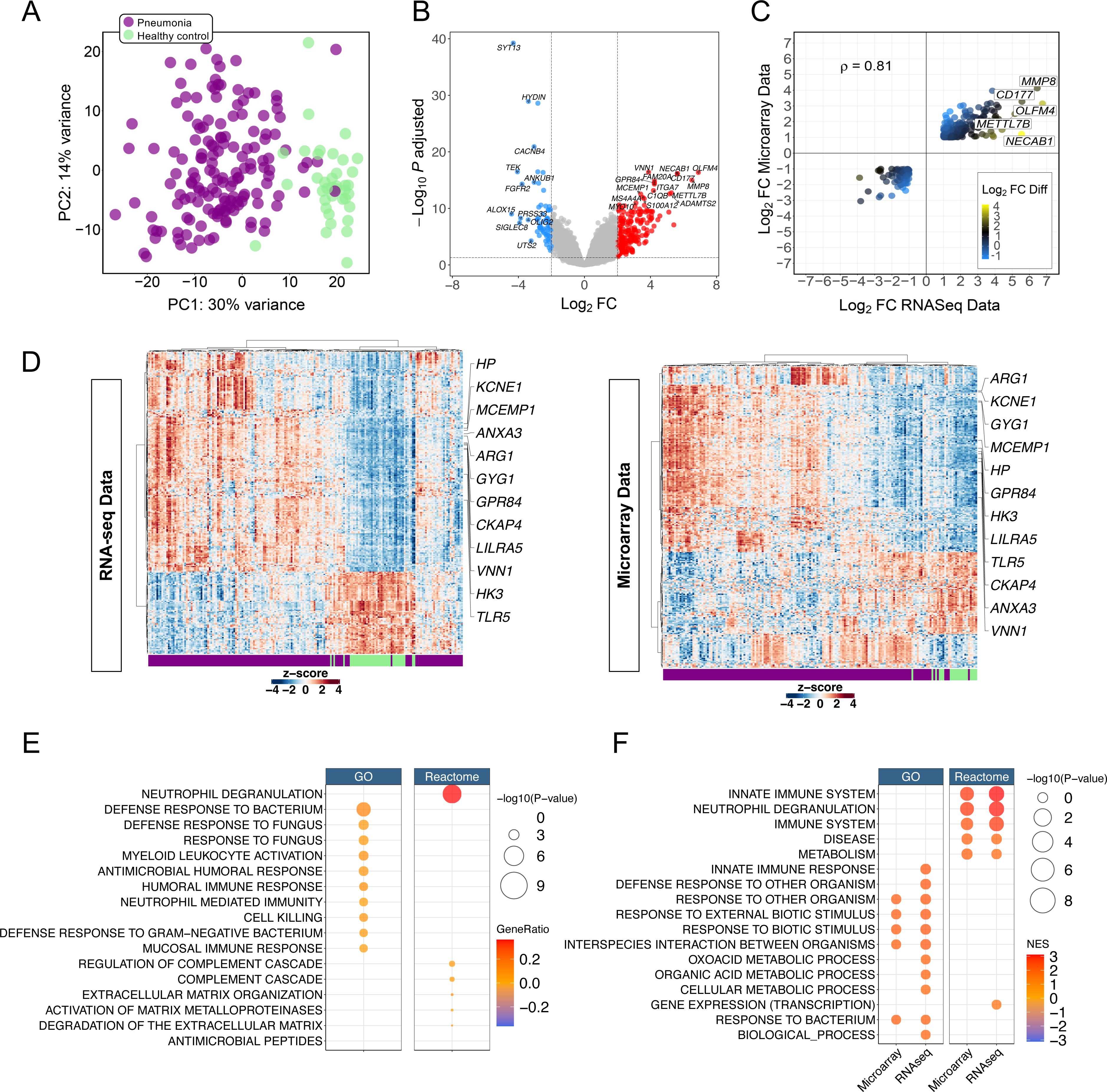
(A) PCA of transcriptome profiles from pneumonia and healthy control samples. Two first principal components (PC1 and PC2) are shown. (B) Volcano plot showing the DEGs between conditions: pneumonia vs. healthy control. Downregulated genes are colored blue and upregulated genes are colored red (thresholds: adjusted *P*-value = 0.05, log_2_FC = |2|). (C) Correlation between log_2_FC of DEGs obtained from the comparison pneumonia vs healthy controls in RNA-seq/Microarray data. Only genes with adjusted *P*-value < 0.05 and log_2_FC > |1| are displayed. The color scale represents the differences between the log_2_FC values of both analyses. The *P*-value of the correlation is 2.2e^−16^ and only names of genes with a Log_2_FC > |5| are shown. (D) Two-way hierarchical clustering analysis heat map of DEGs between pneumonia and healthy control samples in RNA-seq and microarray validation cohorts. Each row represents one transcript; each column represents one patient. The bar at the bottom indicates the sample phenotype. Only genes with a log_2_FC > |1| and adjusted *P-value* < 0.05 were represented in the heatmap and only the genes that where common in the top 40 with the lower *P-value* in both analysis where printed. Expression intensity is indicated by color (Red = high expression, Blue = low expression). (E) Dot plot from ORA pathway analysis of common DEGs between RNA-seq and microarray cohort with adjusted *P*-value (FDR) < 0.05 and a log_2_FC > |1.5| for the comparison pneumonia patients *vs*. Healthy controls using GO and Reactome as reference. Size along the x-axis indicates the number of genes in the input list that are annotated to the corresponding term / number of genes in input list (gene ratio). Dot colors correspond to the different FDR *P*-values associated with the pathways. (F) Dot plot from GSEA pathway analysis of common DEGs between RNA-seq and microarray cohort with FDR *P*-value < 0.05 and a log_2_FC > |1.5| for the comparison pneumonia patients *vs*. Healthy controls using GO and Reactome as reference. Size of the dots correspond to along FDR *P*-values associated with the pathways. Dot colors correspond to the pathway normalized enrichment scores (NES) values.

To prioritize significant changes in gene expression, we applied a selective criterion, including an adjusted *P*-value < 0.05 and a log_2_FC > |2|. This selection process yielded 272 DEGs (**Figure 2B)**, with 208 exhibiting upregulation and 64 genes showing downregulation, indicating a prevailing pattern of overexpression among these genes.

Next, we compared the DEGs identified in our RNAseq cohort, with those inferred from the analysis of the Wallihan et al. study ^68^ (accession number GSE103119). The dataset by Wallihan et al. included transcriptomic data from blood samples of pediatric pneumonia cases (*n* =152) and healthy controls (*n* = 20). Our comparison revealed 1,729 common DEGs (**Table S2**). Focusing on the DEGs with significant expression changes compared to healthy controls (adjusted *P*-value < 0.05 and log_2_FC > |1|; *n* = 310 genes), we found a strong positive correlation (ρ = 0.81; *P*-value < 2.2e^−16^) between log_2_FC values obtained from both datasets (**Table S2**; **Figure 2C**). Furthermore, the expression pattern of these 310 shared genes clearly segregated pneumonia cases from healthy controls, forming two distinct clusters in both our RNA-seq discovery cohort and Wallihan’s microarray dataset (**Figure 2D**).

### Functional enrichment analysis of differentially expressed genes

To further characterize DEGs from a functional point of view, we conducted an over-representation analysis (ORA), and a gene-set enrichment analysis (GSEA) based on gene ontology (GO) and Reactome using only validated DEGs with a positive correlation between the discovery and the validation datasets. We set a threshold of an adjusted *P*-value < 0.05 for significance and a |log_2_FC| > 1.5. The analysis revealed that the most significant pathways (adjusted *P*-value < 0.01) are associated with essential immune processes, primarily related to the innate response to severe CAP (**Table S3**; **Figure 2E**).

The Reactome-ORA analysis indicated a robust association between the selected DEGs, including *MMP8*, *OLFM4* or *CD177*, and an innate response characterized by ‘neutrophil degranulation’. This pathway displayed the highest significance (adjusted *P*-value = 6.5e^−12^); **Table S3**. Consistently, GO-ORA results identified differences in neutrophil-related immune pathways, including ‘leukocyte mediated immunity’ (adjusted *P*-value = 7.4e^−5^), ‘myeloid leukocyte mediated immunity’ (adjusted *P*-value = 0.001), ‘neutrophil mediated immunity’ (adjusted *P*-value = 5.1e^−5^), ‘myeloid leukocyte activation’ (adjusted *P*-value = 1.4e^−5^), ‘granulocyte activation’ (adjusted *P*-value = 0.002), and ‘neutrophil activation’ (adjusted *P*-value = 0.001) (**Table S3**; **Figure 2E**).

Furthermore, there is a significant overlap in enrichment results from GO and Reactome, particularly regarding humoral immunity processes. Notably, GO-ORA identified ‘humoral immune response’ (adjusted *P*-value = 4.4e^−5^), ‘antimicrobial humoral response’ (adjusted *P*-value = 1.4e^−5^) and ‘antimicrobial humoral immune response mediated by antimicrobial peptide’ (adjusted *P*-value = 0.0003) (**Table S3**; **Figure 2E**) as highly significant pathways. Likewise, Reactome-ORA highlighted the involvement of various humoral responses in severe CAP, including the complement system (‘regulation of complement cascade’ [adjusted *P*-value = 0.001] and ‘complement cascade’ [adjusted *P*-value = 0.003]), as well as humoral immunity mediated by ‘antimicrobial peptides’ (adjusted *P*-value = 1.2e^−9^) (**Table S3**; **Figure 2E**).

Most significant pathways from GO-ORA were those implicated in ‘defense response to bacterium’ (adjusted *P*-value = 2.3e^−8^), ‘defense response to fungus’ (adjusted *P*-value = 1.0e^−5^) and related processes (‘defense response to Gram-negative bacterium’ and ‘antibacterial humoral response’) (**Table S3**; **Figure 2E**). It is not surprising that genes related to defense against bacteria were overrepresented in the overall analysis as most of the samples in the dataset correspond to bacterial origin pneumonia.

Additionally, GO-ORA analysis highlighted a significant association of CAP with biological processes that play essential roles not only in innate protection against viral and bacterial invasion but also in disease pathogenesis. This encompasses regulatory pathways related to transcription factors, such as the pro-inflammatory NF-kappaB (’regulation of DNA-binding transcription factor activity’, ‘positive regulation of DNA-binding transcription factor activity’, ‘positive regulation of NF-kappaB transcription factor activity’) as well as pathways indicating inflammatory response (‘regulation of inflammatory response’, ‘positive regulation of inflammatory response’ and ‘acute inflammatory response’) and activation of the cytokine signalling system (‘positive regulation of cytokine production’ and ‘cytokine-mediated signaling pathway’) (**Table S3**; **Figure 2E**).

The results from the GSEA closely mirrored those obtained from both microarray ^68^ and our RNA sequencing cohorts. Remarkably, all differentially regulated pathways exhibited significant over-activation in pneumonia cases (normalized enrichment score [NES] >0) (**Table S3**; **Figure 2F**). The most significant pathways identified in the GO and Reactome analysis overlapped with those detected in the ORA, highlighting key processes such as the ‘response to other organism’ and ‘response to bacterium’ in GO. Additionally, pathways related to the ‘innate immune system’ and processes like ‘neutrophil degranulation’ in Reactome, were also prominently featured.

### Bacterial *vs*. viral pneumonia: diagnostic signature

We conducted an additional PCA analysis to compare the transcriptomic profiles of pneumonia in definitive bacterial (DB) and definitive viral (DV) infections. Our goal was to investigate how these two groups cluster in relation to each other and, in comparison, to the control group. In the first PCA (**Figure 3A**), pneumonias separated distinctly from the control group along PC1, which accounted for 45% of the variation. Specifically, the DB clustered at one pole of the plot, the controls in the opposite side, and the DV profiles in between. A separate PCA including only DB (*n* = 40) and DV (*n* = 9) transcriptomic profiles samples, revealed a relatively weak separation between the two groups. DV profiles primarily clustered to one side along its PC1 (28% of the variation), albeit with some slight mixing with other DB samples; **Figure 3A**.

**Figure 3.**
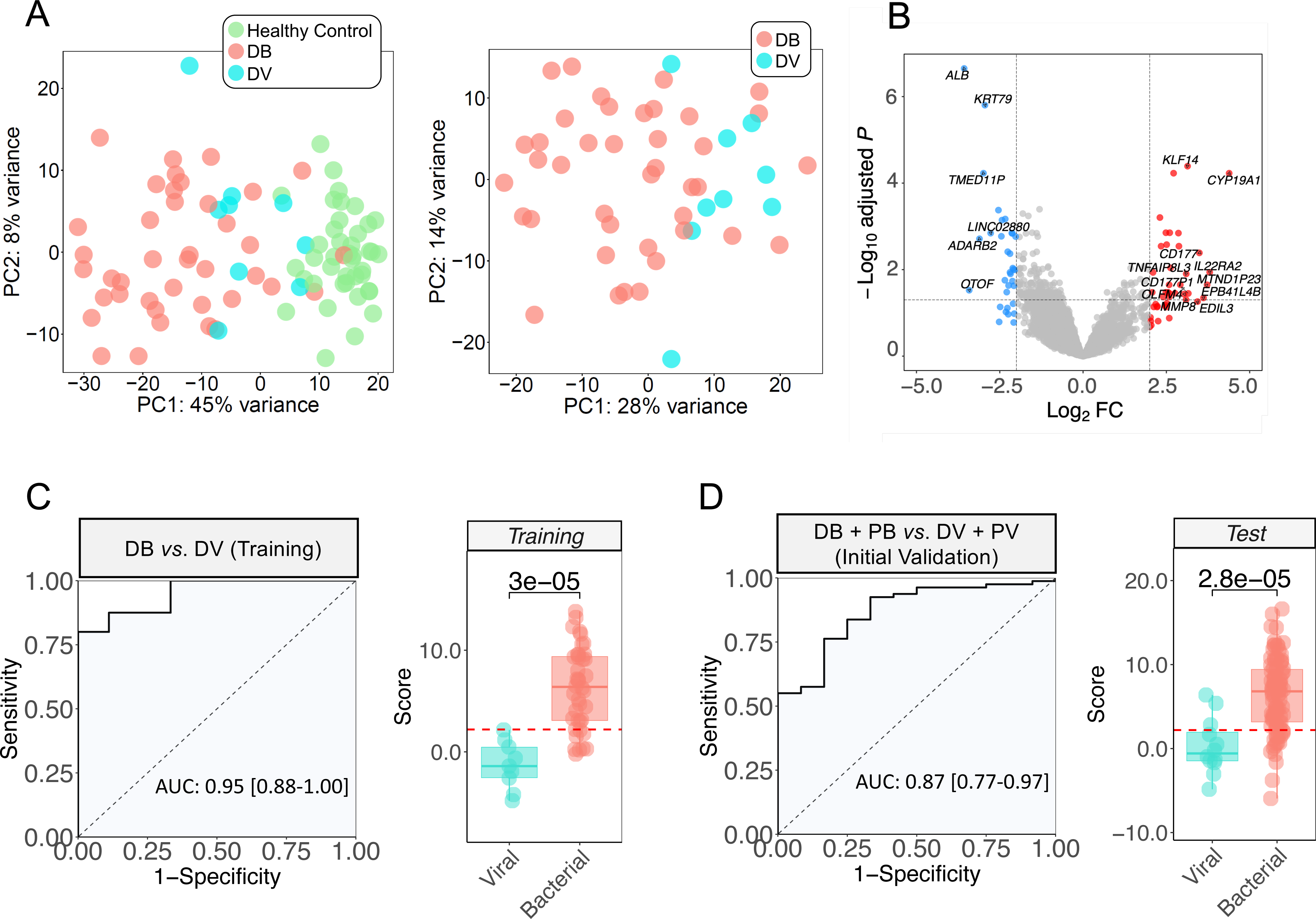
(A) PCA of transcriptome profiles of DV and DB pneumonias and healthy control samples. Two first principal components (PC1 and PC2) are shown. (B) Volcano plot showing the DEGs between conditions: DB pneumonia vs. DV pneumonia (DV). Downregulated genes are colored blue and upregulated genes are colored red (thresholds: adjusted *P*-value = 0.05, log_2_FC = |2|). (C) Receiver operating characteristic curves (ROC) based on the specific 5-transcript signature from the training cohort including the area under the curve (AUC) and 95% confident intervals (CIs) values (left). Boxplots of the predicted values using the optimal model in training cohort. Wilcoxon *P*-value is also displayed (right). (D) Receiver operating characteristic curves (ROC) based on the specific 5-transcript signature from test cohort including the area under the curve (AUC) and 95% confident intervals (CIs) values (left). Boxplots of the predicted values using the optimal model in training cohort. Wilcoxon *P*-value is also displayed (right). Red dashed line represents the optimal cutpoint.

Our differential expression analysis between DB and DV pneumonias identified 282 DEGs (FDR of 5%; **Table S4**). The majority showed under-expression in DB with respect to DV pneumonia (*n* = 217; 77%). However, when considering genes with more substantial expression changes (setting a threshold of adjusted *P*-value < 0.05 and a |log_2_FC| > 2), we found that the proportion of over-expressed genes in DB pneumonia (*n* = 30; 55%) was slightly higher than the proportion of under-expressed genes (*n* = 25; 45%); **Table S4; Figure 3B**. Interestingly, *ALB* was the gene yielding the lowest adjusted *P*-value (1.88e-7) in the comparison between DB and DV patients (**Table S4**; **Figure 3B; Figure S1**).

To further investigate, we selected a sub-set of DEGs candidates from the DV vs. DB differential expression in pneumonia samples (*n* = 36; see Material and Methods). We then employed the LASSO regression algorithm to identify the optimal transcriptomic signature for distinguishing between these two types of infection.

The best model for differentiation between viral and bacterial pneumonia comprised five genes (*FAM20A*, *BAG3*, *TDRD9*, *MXRA7* and *KLF14*), forming the optimal signature (**Table S5**). Among these genes, four were over-expressed (*FAM20A*, *TDRD9*, *MXRA7* and *KLF14*) while one (*BAG3*) showed under-expression in bacterial compared to viral pneumonia. This transcriptomic signature statistically discriminated between viral and bacterial pneumonia with high accuracy (*P*-value = 3.0e^−5^; optimal cut-off value = 2.204; **Figure 3C)** and demonstrated excellent performance in the training set, generating an AUC value of 0.95 [0.88-1.00] (sensitivity = 0.80 and specificity = 1.00) (**Figure 3C; Table S6**). Moreover, the signature reliably differentiated bacterial pneumonia from healthy controls (Area under the curve [AUC]: 0.91 [0.84-0.98]) (**Figure S2**).

Due to the limited number of samples classified as viral pneumonia (DV = 9; probable viral [PV] = 3), we were unable to split the dataset into training and test sets conventionally. Instead, we adopted an initial validation approach, evaluating the performance of the transcriptomic signature in a test dataset that included not only the samples confidentially classified as DB and DV, which had been already used in the training dataset for the signature discovery (DB = 40; DV = 9), but also all probable bacterial (PB) (n = 58) and PV (n = 3) pneumonias (DB + PB = 98; DV + PV = 12; **Figure 1**).This 5-transcripts signature identified in the training cohort exhibited robust discriminatory capacity when validated in this test set (*P*-value = 2.8e^−5^, **Figure 3D**). In this scenario, the ROC curve also indicates high accuracy, resulting in an AUC value of 0.87 [0.77–0.97] (sensitivity = 0.76 and specificity = 0.83); **Figure 3D**; **Table S6**. Considering only PV and PB samples we obtained an expected decrease in the AUC value (0.69 [0.40-0.97]), as these samples are coded with a phenotype of lower clinical confidence, and are therefore more likely to contain viral samples within the PB set, bacterial samples within the PV set, or mixed infections (see score values for classification in **Figure 3C** and **3D**; see also **Figure S3A)**.

The performance of our 5-transcriptomic signature was also compared to the previously described 2-transcript signature that generically differentiate viral from bacterial infections in febrile children using *IFI44L* and *FAM89A* transcripts ^30^. Although the 2-transcript signature showed a good performance discriminating viral and bacterial pneumonia (AUC: 0.87 [0.76-0.96]) in our discovery dataset, the AUC value was lower than that obtained from the specific 5-transcript signature we propose; **Figure S3B.** When we tested the 2-transcripts viral/bacterial signature in the PB vs. PV subset, the AUC value was also lower than the AUC from the pneumonia signature (AUC:0.44 [0.00–0.89]); **Figure S3C**. Finally, to check the best performance that we could achieve with these two transcripts in our discovery dataset, we re-trained the model using *IFI44L* and *FAM89A* transcripts. In this case we obtained an AUC value of 0.88 [0.77-0.98]; **Figure S3D**, which is still below the AUC from the specific pneumonia signature (AUC: 0.95 [0.88–1.00]).

Additionally, we tested the 5-transcripts signature and the 2-transcript signature from Herberg et al. ^30^ in the bacterial and viral pneumonia samples (excluding co-infections) from dataset of Wallihan et al. ^68^. Both signatures yielded AUC values close to 0.5 for this dataset, with the 5-transcript signature achieving and AUC of 0.52 [0.40-0.64], and the 2-transcripts signature from Herberg et al. reaching an AUC of 0.56 [0.45-0.68]. These results indicate limited discrimination ability for these signatures, within this specific dataset ^30^.

### Validation of the 5-transcripts pneumonia’s signature

We validated the diagnostic accuracy of the proposed signature using new gene expression data obtained from an independent cohort of children with pneumonia (**Table 2**, **Table S7**). The validation results confirm that the 5-transcript signature can accurately differentiate between definitive viral and bacterial pneumonias in this new cohort, achieving an impressive AUC of 0.92 [0.83-1.00]; **Figure 4A**, **Figure 4B**.

**Figure 4.**
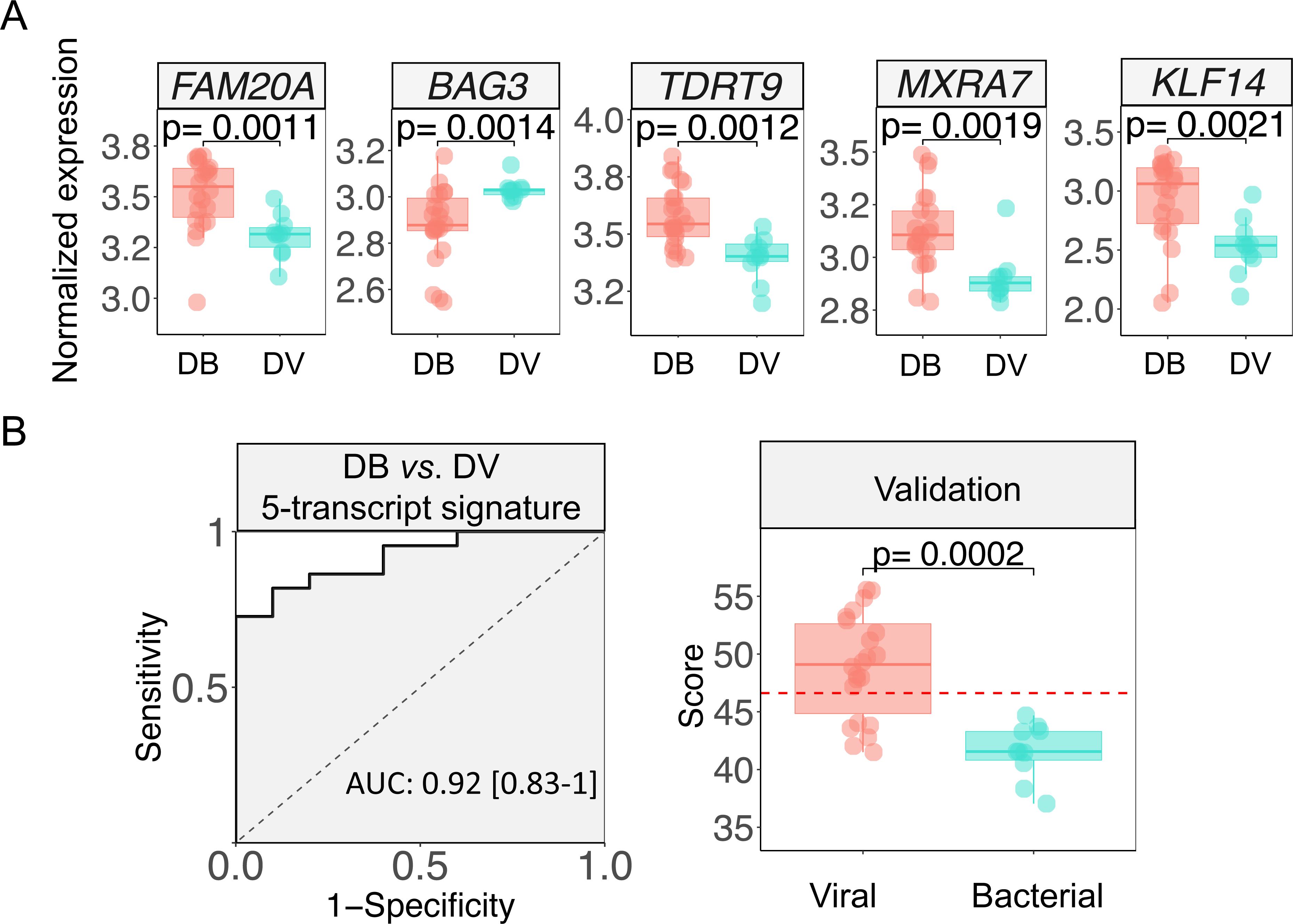
(A) Boxplots showing the expression values of the genes included in the 5-transcript signature in the validation cohort. (B) ROC curve, AUC value with confidence interval and boxplot of the predicted value obtained from applying the 5-transcript viral/bacterial signature coefficients in the validation cohort. Wilcoxon test P-values are displayed in the boxplots.

**Table 2.** Clinical features of the validation cohort. Wilcoxon and Fisher exact test were used to assess statistical significance between groups in numerical and categorical variables respectively IV: invasive ventilation; NIV: non-invasive ventilation.

|  | Healthy controls | DB vs. DV |  |  |  |  |
| --- | --- | --- | --- | --- | --- | --- |
|  | Controls<br>(n = 35) | DB<br>(n = 22) | n | DV<br>(n= 10) | n | P-value |
| **Gender (Male)** | 54.3% (19/35) | 63.6% (14/22) | 22 | 60% (6/10) | 10 | 1 |
| **Age (years)** | 4.2 (1.24-6.17) | 4.8 (0.1 - 17.5) | 22 | 2.4 (0.1 - 15.6) | 10 | 0.167 |
| **Days hospitalized** | – | 14.0 (3.5-14) | 22 | 6 (4.75-8.5) | 8 | 0.038 |
| **PICU admission** | – | 54.5% (12/22) | 22 | 40% (4/10) | 10 | 0.703 |
| **PICU days** | – | 2.0 (0.0 - 14.0) | 22 | 7,5 (2.00, 8.00) | 4 | 0.843 |
| **Death** | – | 4,5% (1/22) | 22 | 0% (0/9) | 10 | 1 |
| **Treatments** |  |  |  |  |  |  |
| *Inotropes* | – | 32% (7/22) | 22 | 20% (2/10) | 2 | 0.379 |
| *IV* | – | 31.8% (7/22) | 22 | 30% (3/10) | 9 | 0.750 |
| *NIV* | – | 13,6% (3/22) | 22 | 30% (3/10) | 9 | 1 |
| *Oxygen* | – | 54.5% (12/22) | 22 | 60% (6/10) | 9 | 0.637 |
| **Analytical parameters** |  |  |  |  |  |  |
| *CRP (mg/L)* | – | 208.4 (46.2 - 375.4) | 16 | 18.2 (1.3 - 52.5) | 10 | 0.084 |
| *Platelets (10<sup>9</sup>/L)* | – | 249.0 (17.0 - 1299.0) | 19 | 271.5 (141.0 - 522.0) | 10 | 0.449 |
| *White Cells (10<sup>9</sup>/L)* | – | 13.5 (4.8 - 28.3) | 19 | 5.8 (2.8 - 11.5) | 10 | 0.001* |
| *Lactate (mmol/L)* | – | 1.4 (0.6 - 5.9) | 8 | 0.7 (0.5 - 1.7) | 4 | 0.147 |
| *Neutrophils (10<sup>9</sup>/L)* | – | 9.6 (3.7 - 24.0) | 15 | 3.0 (0.9 - 5.1) | 9 | 0.001* |
| *Monocytes (10<sup>9</sup>/L)* | – | 0.8 (0.1 - 3.4) | 15 | 0.4 (0.1 - 2.0) | 8 | 0.258 |
| *Lymphocytes (10<sup>9</sup>/L)* | – | 2.5 (0.2 - 6.0) | 15 | 2.6 (0.9 - 4.7) | 9 | 0.765 |

To ensure that the effectiveness of the signature was not influenced by disease severity or specific pathogens, we conducted a stratified analysis based on the causal pathogens of definitive bacterial (DB) pneumonias—*S. pneumoniae*, *S. aureus*, and *S. pyogenes*—and by severity (mild/moderate *vs*. severe). The results affirmed the robustness of the signature across different bacterial pathogens: AUC values of 0.95 [0.87-1.00] for *S. pneumoniae*, 0.82 [0.54-1.00] for *S. aureus*, and a perfect AUC of 1 [1.00-1.00] for *S. pyogenes* (**Figure 5**). Moreover, the signature consistently demonstrated high performance in distinguishing between viral and bacterial pneumonias across both mild/moderate and severe cases, with AUC values of 1 [1.00-1.00] for mild/moderate cases and 0.92 [0.77-1.00] for severe cases (**Figure 5**).

**Figure 5.**
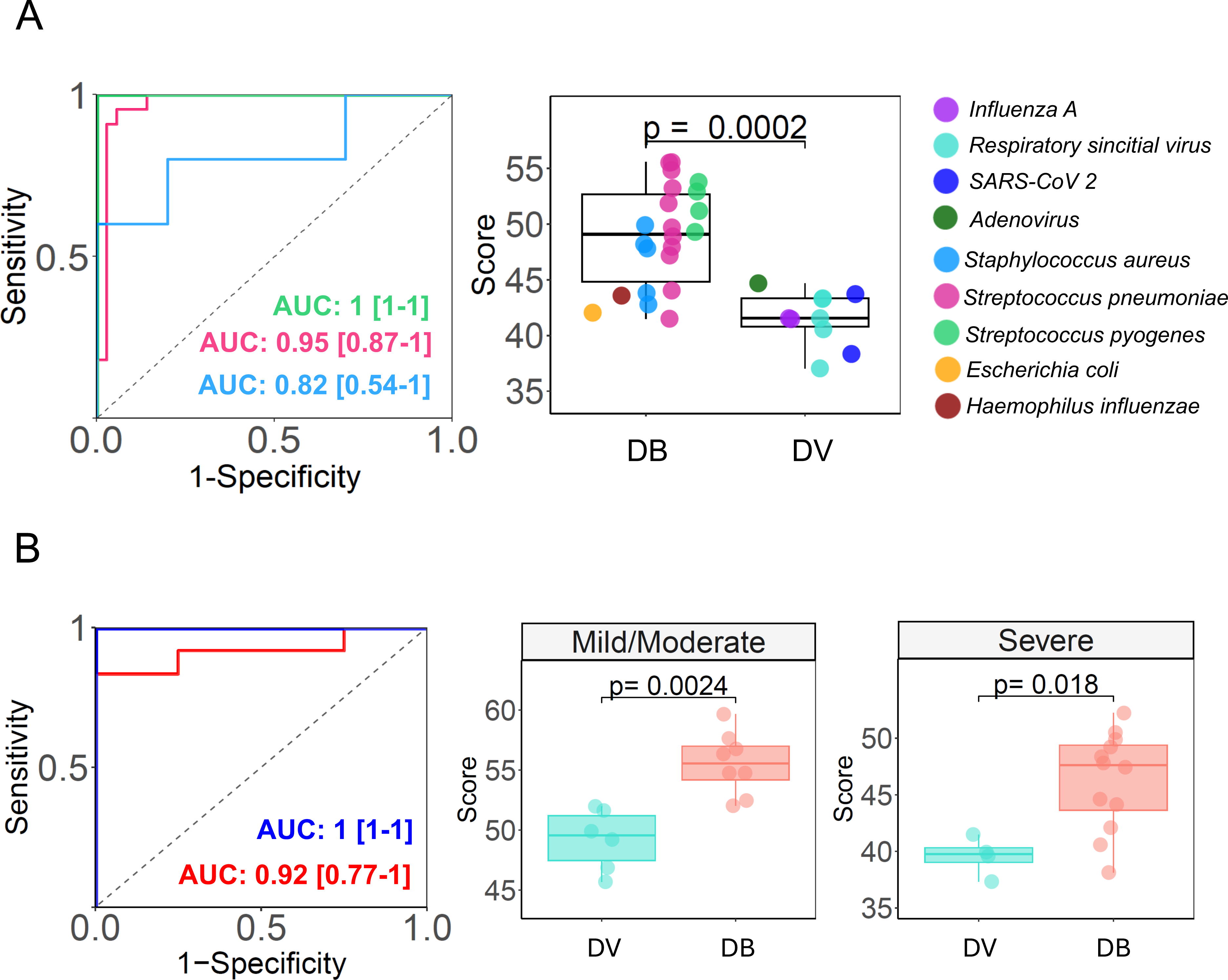
(A) ROC curves, AUC values and boxplots of the performance of the 5-transcript signature in pneumonias with different causal pathogens. Only bacterial groups with more than 3 samples were considered for the AUC and ROC analysis. (B) ROC curves, AUC values and boxplots of the performance of the 5-transcript signature in pneumonias with different severities (severe or mild/moderate).

### Co-expression analysis of viral *vs*. bacterial pneumonia

To understand the distinct host mechanisms and key genes involved in the specific response to viral and bacterial pneumonia, we conducted a co-expression network analysis using the sub-set of pneumonia samples with DB or DV origin. The analysis detected 16 modules of co-expressed genes (**Figure S4**), with three of them significantly correlated (*P*-value < 0.05) with viral/bacterial phenotypes (**Figure 6A**; **Figure 6B**). The blue module ([EVL module], *R* = −0.4; *P*-value = 0.0043; 661 genes) and darked module ([TGFBR3 module], *R* = −0.32; *P*-value = 0.025; 73 genes) exhibited negative correlation with bacterial pneumonia. In contrast, the salmon module [MAPK14] showed a positive correlation (*R* = 0.32; *P*-value = 0.026; 1,132 genes) with bacterial pneumonia (**Figure 6A**; **Figure 6B**). The EVL and TGFBR3 modules clustered together on the dendrogram, indicating their involvement in similar biological processes globally activated during viral pneumonia **(Figure 6A)**. Genes within these significant modules displayed a strong correlation between trait significance (GS) and module membership (MM), suggesting that highly interconnected genes (higher MM) within the module are closely related to the causal pathogen of pneumonia (**Figure 6B**). These driver genes, located in the upper right side of the plots, are pivotal for predicting viral or bacterial pneumonia.

**Figure 6.**
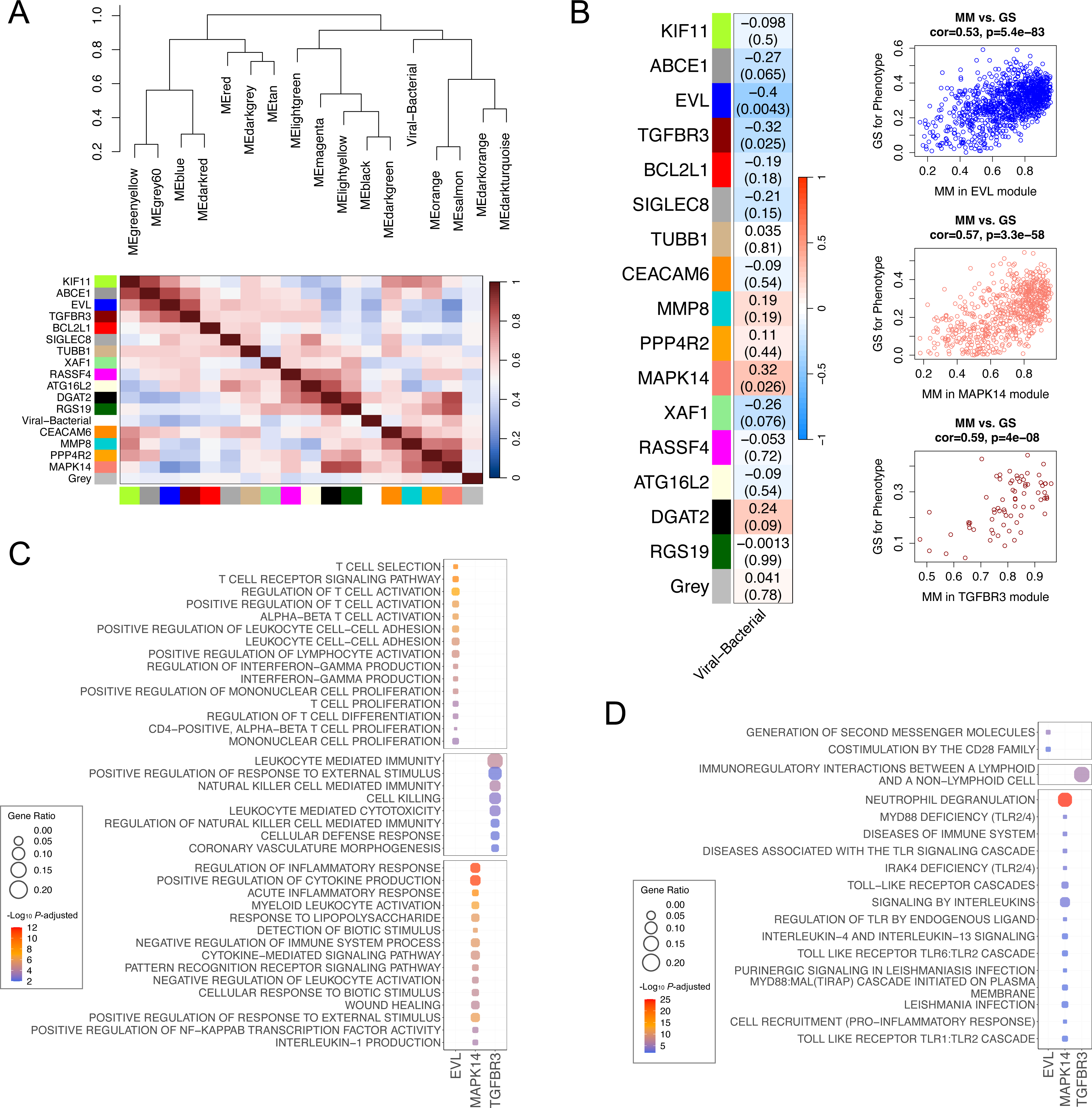
(A) Hierarchical clustering eigengene dendrogram and heatmap showing relationships among the modules and pneumonia etiology (DB and DV pneumonias). Gene names on the left of the heatmap are the hub genes of each module. (B) Correlation values heatmap between co-expression modules and DB and DV conditions (DV was considered the reference group). Upper value shows to individual correlation value of the module with musical stimuli (left). Correlation values are also indicated with different colors (legend gradient color bar). *P*-values of these correlations are represented in brackets (lower values). Plots showing comparison between MM (module membership) and GS (correlation with bacterial/viral condition) of genes from the most significant modules detected. (C) Top 15 statistically associated GO biological processes detected for each of the significantly correlated modules (D) Top 15 statistically associated Reactome pathways detected for each of the significantly correlated modules

Functional assessment of the significant modules unveiled different immune responses for viral and bacterial pneumonia. The modules over-regulated in viral pneumonia showed enrichment in terms associated with both innate and adaptative immune responses. For example, the EVL module was linked to adaptative T-cell response (T-cell selection, T-cell receptor signalling, T-cell activation, differentiation, and proliferation) and interferon production (interferon gamma production) (**Figure 6C; Table S8**), while the TGBR3 module participated in ‘natural killer mediated immunity’ (innate response) and, as in the case of the EVL module, in an adaptative response involving ‘leukocyte mediated cytotoxicity’ and ‘T-cell mediated immunity’ (**Table S8**).

Conversely, the significant pathways related to genes from the bacterial MAPK14 module primarily represent a different innate response characterized by ‘positive regulation of cytokine production’, ‘cytokine-mediated signalling pathway’, ‘acute inflammatory response’, ‘pattern recognition receptor signalling’ and ‘myeloid leukocytes activation’ (Granulocytes, monocytes, macrophages, and dendritic cells). These findings were further supported by an over-representation analysis using the Reactome database, emphasizing the role of MAPK14 module genes in innate immune responses, including neutrophil degranulation, various toll-like receptor (TLR) cascades, and signalling by interleukins (**Figure 6D; Table S8**).

## Discussion

Pneumonia is a complex disease characterized by diversity in its etiology, symptomatology, and clinical management. This complexity extends to decisions regarding the prescription of antibiotics and hospitalization for children, which remains a significant issue in terms of medical cost ^13^. Some efforts have been made to identifying molecular biomarkers to assist in diagnosing and predicting pneumonia outcomes. However, there is still not an optimal test to accurately diagnose pneumonia in children or to distinguish between bacterial and viral pneumonia. Consequently, antibiotics are often prescribed when a bacterial infection cannot be ruled out leading to unnecessary cost, and contributes to the development of antimicrobial resistance ^9,69^.

Our current analysis reveals that pneumonia in children induce extensive modifications in the host’s transcriptome. Our findings indicate that pneumonia alters the expression of more than 5,000 genes in blood of affected children. Subsequent validation of these differential expression results in an independent cohort of pediatric pneumonia cases confirmed these findings. Additionally, two independent enrichment approaches (ORA and GSEA) using GO and Reactome as reference databases revealed a convergence of altered pathways within the immune system. Specifically, these pathways involve the activation and migration of leukocytes, with significant focus on neutrophil transcripts. During pneumonia, there is recruitment of neutrophils into the lungs. In cases of acute lung injury, a distinct pattern of activated neutrophils, referred to as the neutrophil activation signature ^70^, emerges. This pattern is unique to the process of neutrophils migrating towards infected lung tissue, leading to various responses, including the release of bactericidal enzymes through degranulation, the discharge of cytokines and reactive oxygen species, and the generation of neutrophil extracellular traps. While these activities facilitate the elimination of causative pathogens, they may also result in tissue damage and exacerbate acute inflammation ^71–73^.

Most prominent over-expression changes detected in our RNA-seq cohort, which were subsequently validated in an independent dataset, were represented by genes that produce proteins secreted by secondary granules released from neutrophils. These genes include *OLFM4*, *MMP8*, and *CD177*, and some of them have been previously reported as candidate biomarkers for the diagnosis/prognosis of CAP and predictors of sepsis ^74–76^. Matrix Metalloproteinase-8 (MMP8) is a collagenase derived from neutrophils that has implications in a wide variety of pulmonary pathologies. It can degrade all components of the extracellular matrix, and plays a role in neutrophil migration ^77^. Its expression has already been shown to be significantly higher in pneumonia patients compared to healthy controls, representing one of the most over-expressed in adults pneumonia samples from various studies ^27,78^, and also in our pediatric cohort. Serum and plasma levels of MMP8 protein have been proposed for diagnosing CAP, and as prognostic biomarker of fatal outcome in septic patients and predictive of serious bacterial infection, suggesting that could be an inflammatory modulator in sepsis ^79–81^. However, its poor specificity limits its use as a single protein diagnostic biomarker. Our findings support the evidence that *MMP8* gene is also over-expressed in children with pneumonia. Similarly, we observed a significant over-expression of the *OLFM4* gene in pneumonia patients. *OLFM4* is a matrix glycoprotein of neutrophil-specific granules and has been described as a marker of severity in infectious diseases. In children, it has been identified as an independent risk factor for poor outcomes in sepsis ^82,83^, and its inhibition has been proposed as a possible therapeutic approach in infected patients (55). *CD177* is a glycosylphosphatidylinositol (GPI)-anchored protein that plays a crucial role in regulating neutrophils by modulating their migration and activation. *CD177* has been identified as the most dysregulated molecule in purified neutrophils from patients with septic shock and severe influenza infection ^84^. Recently, it has also been associated with worse outcome and higher mortality in patients infected with SARS-CoV-2 ^85^ and included in a diagnostic four-gene signature for pediatric sepsis ^74^.

*ABL* was the gene yielding the lowest significant value when comparing DB and DV patients. *ALB* encodes for the albumin protein, the most abundant protein in human blood, but it is primarily expressed in the liver and pancreas (https://www.gtexportal.org/). Although the overall expression of ALB in our blood samples was low (median log_2_ values of 2.07, 5, and 3.09 for DB, DV, and HC, respectively), it showed a significant overexpression in DV compared to DB patients. Some studies have showed that decreased levels of albumin in serum and plasma could be indicative of higher risk of ICU admission and death in sepsis ^86,87^, function as a diagnostic biomarker for infectious diseases ^88^, and a risk factor of a more severe clinical manifestations in COVID-19 patients ^89,90^.

In addition to the neutrophil-related innate response, our analysis identified a common enrichment of terms related to humoral immunity processes, consistent with previous results from pediatric pneumonia cases ^91^, as well as an enrichment of innate processes commonly activated in response to viral and bacterial infections, such as the cytokines release and inflammation. At the same time, however, the over-activation of these processes can also be responsible for the clinical consequences of severe pneumonia, increasing the risk of severe sepsis and death ^92,93^.

Since pneumonia can be caused by a broad range of viral or bacterial pathogens, it is essential to investigate new approaches to differentiate the pathogenic origin of the disease, which may support decisions regarding antibiotic prescriptions. Molecular methods for detecting causative bacteria remain less effective in pneumonia patients, mainly due to challenges in obtaining samples from the infection site and the frequent absence of positive results from accessible sites such as blood ^94^. Moreover, detecting viruses in nasopharyngeal secretions from pneumonia patients is common due to colonization, but this does not exclude the possibility of a concurrent bacterial infection ^95,96^. A comparison of gene expression between DV *vs*. DB pneumonia revealed 282 DEGs between both categories. Functional studies of the co-expression modules driving the host specific responses to viral or bacterial pneumonia highlighted differentially activated immunological pathways in both pneumonia etiologies. We identified two significant modules associated with viral pneumonia (EVL and TGBR3) and one associated with bacterial pneumonia (MAPK14). Genes from the MAPK14 module are primarily involved in innate responses related to cytokine production, acute inflammation, neutrophil degranulation, deficiencies in TLR cascade and interleukin signalling. TLR receptors are critical in constraining the proliferation and dissemination of invading bacteria. Activation of TLRs triggers innate immune signalling pathways and transcriptional upregulation of proinflammatory mediators that recruit neutrophils to pulmonary tissue during bacterial infection ^97^. An effective and timely inflammatory response is key to pathogen clearance and control the infection in pneumonia. However, an exacerbated and prolonged release of pro- and anti-inflammatory cytokines, followed by neutrophil hyper-responsiveness, is associated with an increased risk of severe sepsis and poor outcomes in CAP ^92,98^. TLRs can detect various conserved patterns from both viral and bacterial pathogens. Interestingly, significant pathways involving TLRs detected in the bacterial module are among the TLRs cascades commonly activated by bacterial pathogen-associated molecular patterns (PAMPs; e.g. lipoproteins TLR1/TLR2/TLR6 and lipopolysaccharide TLR4) ^99^. Most TLRs, except TLR3, signal *via* myeloid differentiation primary response 88 (MyD88) and interleukin-1 receptor-associated kinase 4 (IRAK-4). IRAK4 is critical for MyD88-dependent TLR signaling, and patients with IRAK4 mutations are susceptible to recurrent bacterial infections ^100,101^. Patients with both IRAK-4 and MyD88 deficiencies are predisposed to severe bacterial infection ^102^ and defects in MyD88-mediated TLR signalling make children much more susceptible to pneumonia ^103^. The canonical TLR pathway can activate NF-κB and MAPKs through different mediators; in this regard, the hub gene of our bacterial module, *MAPK14,* is a key gene in the development of ventilator-associated bacterial pneumonia in adults ^104^.

Collectively, these findings suggest that immune responses to severe bacterial pneumonia are primarily characterized by dysfunction in the innate TLR signalling. This dysfunction leads to an exacerbated inflammatory response, driven by cytokine release and neutrophil degranulation, which may play a pivotal role in the development of the severe clinical symptoms observed in these patients.

In the case of viral pneumonia-correlated modules (EVL and TGBR3), we identified a significant representation of adaptative immunity mediated by T-cells. This includes processes such as T-cell selection, T-cell receptor signalling, T-cell activation, differentiation, and proliferation, especially genes within the EVL module. The TGFBR3 module also reveals pathways related to adaptative immunity, such as “leukocyte mediated cytotoxicity” and “T-cell mediated immunity”. T-cells are essential in combating viral infections, by inducing cell death through cytokines release ^105^. Notably, both CD4+ and CD8+T-cell increase in population during the first days to weeks of an acute viral infection ^106^. CD8+ cells have been described to be critical in clearing various viral infections in mouse models ^107^, while CD4+ T-cells serve as helpers for B-cells and CD8+ T-cells, also possessing independent antiviral function through cytokine secretion ^108^. Of interest, polymorphisms in the hub gene *TGFBR3* were previously associated with a history of pneumonia in Sickle Cell Disease patients ^109^. EVL expression was associated with poor prognosis in sepsis ^110^.

We have identified a 5-transcript blood signature that can accurately differentiate bacterial from viral pneumonias with an AUC of 0.95 (sensitivity = 0.8; specificity = 1.0), demonstrating superior discriminant power between viral and bacterial pneumonia compared with previously published non syndrome specific signatures for differentiating viral from bacterial infections in children (AUC: 0.87 [0.75-0.96] and 0.88 [0.78-0.98] in the re-trained model) ^30^. However, as the general signature also performs effectively in this cohort, both methods could be implemented complementarily in a clinical context for the definitive confirmation of the diagnosis. The 5-transcript signature’s performance remains high when including pneumonia samples of both PB and PV origin (AUC = 0.87; sensitivity = 0.76; specificity = 0.83), affirming the robustness and discriminatory power of the transcriptomic signature. Among the genes selected in this signature, some are associated with anti-viral or bacterial activity. For instance, *BAG3* is a cochaperone recently linked to intracellular bacterial proliferation ^111^*. FAM20A*, a pseudokinase, exhibits increased expression in lung and liver ^112^, in sepsis ^113^, and in blood neutrophils of acute respiratory distress syndrome patients ^70^. The *TDRD9* gene was previously described in a transcriptomic signature for sepsis derived from CAP ^114^. *MXRA7* is highly expressed in murine and human ocular tissues and may play a role in pathological processes involving injury, neovascularization, and wound healing, although its function remains mainly unknown ^115^. Finally, the Krüppel-like transcription factor *KLF14* exhibits a modulatory role of in macrophages-driven inflammation ^116^, and has been found to be up-regulated in septic patients ^117^. Its potential regulatory function in sepsis positions this gene as a therapeutic target candidate for sepsis.

We have also observed that the 5-transcript signature identified in the present study and the signature identified by Herberg et al. ^30^ for differentiating viral from bacterial infection did not prove effective in the pneumonia cohort from Wallihan et al. ^68^. This outcome can be explained by the fact that the vast majority of patients with bacterial pneumonia in the Wallihan et al. cohort have infections caused by *Mycoplasma pneumoniae* (30/35; 86%). These results suggest that the host gene expression response to this pathogen is specific and distinct from that triggered by other pneumonia-causing bacteria. The fact that *M. pneumoniae* is generating worrying outbreaks in the post-COVID-19 pandemic era ^118,119^ deserves further dedicated investigation.

The validation of the transcriptomic signature in an independent cohort of children with pneumonia further reinforces its high specificity and sensitivity in accurately distinguishing bacterial from viral infections. Importantly, our analysis confirmed that the effectiveness of the signature is independent of both the causal bacterial pathogen responsible and the severity of the disease. This robust performance across diverse clinical contexts underscores the potential of this signature as a reliable diagnostic tool for pediatric pneumonia, offering a significant advancement in precision and reliability over current diagnostic methods.

Our study involved hospitals from different countries, reducing the potential bias from specific populations or areas. It has characterized and independently validated transcriptomic changes caused by CAP in hospitalized children. The primary limitation of our study is the limited number of samples from children with viral pneumonia. Therefore, further validation is required, utilizing additional datasets that encompass well-defined pneumonia infections of both viral and bacterial origins. Despite this limitation, our results reaffirm the classification of pediatric patients into viral and bacterial pneumonia categories based on the clinical algorithm developed by Herberg et al. ^30^.

We have uncovered various immune mechanisms and genes that are distinctly activated in viral and bacterial pneumonia, making a significant advance in the field. Most notably, our 5-transcript signature stands out for its ability to accurately differentiate between bacterial and viral pneumonia, even in cases of uncertain origin. Crucially, this signature might make such distinctions before the full clinical symptoms emerge. This early diagnostic capability holds the potential to drastically reduce the time to a definitive diagnosis compared to traditional culture-based methods, ultimately shortening hospital stays, and mitigate the consequences of delayed or missed diagnoses, such as unnecessary hospital investigations and antibiotic prescriptions when a bacterial infection is suspected. These benefits position our findings as a major step forward in improving pneumonia management.

## Legends to Supplementary Figures

**Figure S1**. ALB gene expression in virus, bacterial and healthy controls, and *P*-values.

**Figure S2**. ROC curve, AUC value with confidence interval and boxplot of the predicted value obtained after testing the performance of the 5-transcript viral/bacterial signature in differentiating DB from the healthy control group. Wilcoxon test P-values are displayed in the boxplots.

**Figure S3.** (A) Receiver operating characteristic curves (ROC) based on the specific 5-transcript signature from PV and PB pneumonia samples including the area under the curve (AUC) and 95% confident intervals (CIs) values. (B) Receiver operating characteristic curves (ROC) based on the specific viral/bacterial 2-transcript signature (*IFI44L* and *FAM89A*) from DV and DB pneumonia samples including the area under the curve (AUC) and 95% confident intervals (CIs) values. (C) Receiver operating characteristic curves (ROC) based on the viral/bacterial 2-transcript signature (*IFI44L* and *FAM89A*) from PV and PB pneumonia samples including the area under the curve (AUC) and 95% confident intervals (CIs) values. (D) Receiver operating characteristic curves (ROC) based on the re-trained the model using specific viral/bacterial 2-transcript signature (*IFI44L* and *FAM89A*) from DV and DB pneumonia samples including the area under the curve (AUC) and 95% confident intervals (CIs) values.

**Figure S4**. Clustering dendrogram of genes and co-expression modules detected represented by different colors.

**Figure S5**. Soft-thresholding power estimation in DB and viral cohort. Plots represent correlation between soft-thresholding powers and both the scale-free fit index and the mean connectivity.

## Legends to Supplementary Tables

**Table S1.** Differentially expressed genes between pneumonia patients and healthy controls in blood samples (adjusted *P*-value < 0.05).

**Table S2.** Differentially expressed genes between pneumonia patients and healthy controls in the microarray dataset (GSE103119) (adjusted P-value < 0.05). Common differentially expressed genes between the RNA-seq cohort and the microarray cohort (only genes with adjusted *P*-value < 0.05 and log_2_FC > |1|).

**Table S3**. Pathway analysis results obtained from ORA (Over-representation analysis) and GO: Gene Ontology database; BP: Biological Process. Pathway analysis results obtained from ORA (Over-representation analysis) and Reactome database. Pathway analysis results obtained from GSEA (gene-set enrichment analysis) and GO: Gene Ontology database; BP: Biological Process; NES: Normalized enrichment score. Pathway analysis results obtained from GSEA (gene-set enrichment analysis) and Reactome database; NES: Normalized enrichment score.

**Table S4**. Differentially expressed genes between DV and DB pneumonia patients in blood samples (adjusted *P*-value < 0.05).

**Table S5**. Regression coefficients for the 5-transcript signature for differentiating viral from bacterial pneumonia.

**Table S6**. Area under de curve (AUC) with confidence intervals (CI), sensitivity and specificity values for the 5-transcripts signature in training and validation cohorts and for the 2-transcript signature.

**Table S7**. Details on dataset availability.

**Table S8**. Gene Ontology (GO) enrichment analysis obtained from the statistically significant co-expression modules. Reactome enrichment analysis obtained from the statistically significant co-expression modules.

## STAR Methods

### Experimental model and study participant details

This study utilized blood samples from paediatric patients recruited in the European Union Childhood Life-threatening Infectious Diseases Study (EUCLIDS-https://www.euclids-project.eu/) ^53^. The EUCLIDS study enrolled children and young adolescents aged 1 month to 18 years who were admitted to hospitals with suspected sepsis or severe focal infection. The study encompassed a network of 194 hospitals across nine European countries, as well as two hospitals located in The Gambia, Africa. Patient recruitment occurred between July 1, 2012, and December 31, 2015.

Pneumonia phenotype was defined as clinical symptoms compatible with acute respiratory infection and inflammation of one or both lungs with lobar or segmental or multilobar collapse/consolidation on chest X-ray. More specifically, following radiological findings of consolidation/pleural effusion must be met: alveolar consolidation (defined as a dense or fluffy opacity that occupies a portion or whole of a lobe or of the entire lung that may or may not contain air-bronchograms) or pleural effusion (defined as fluid in the lateral pleural space and not just in the minor or oblique fissure) that was spatially associated with a pulmonary parenchymal infiltrate (including other infiltrate) or obliterated enough of the hemithorax to obscure an opacity. It does not include just perihilar consolidation or patchy consolidation. Children with pneumonia (*n* = 154) and age and sex matched healthy controls (*n* = 38; **Table 1**) were selected from the EUCLIDS database ^54^ for transcriptome analysis in whole blood samples collected in PAXgene^TM^ and Tempus™ RNA tubes. In a subsequent analysis, pneumonia samples were further classified using the algorithm developed by Herberg et al. ^30^. C-reactive protein (CRP) and neutrophil threshold criteria were employed where available, with 72.5% and 33.3% of missing CRP values in definitive bacterial (DB) and definitive viral (DV), respectively (see ^30^ for details on clinical characteristics of the cohort and definitions on DB and DV).Thus, DB (*n* = 40) and DV (*n* = 9) pneumonias were selected to study the differences in transcriptome depending on the etiology of the infection. Pneumonias categorized as Probable Bacterial (PB) and Probable Viral (PV) were excluded from this specific analysis to minimize possible confounding factors in stablishing a transcriptomic signature for differentiating bacterial from viral pneumonia in children but were used post-hoc to test the signature in a more realistic clinical scenario (**Figure 1**).

In order to validate the proposed signature, a new independent cohort comprising blood samples obtained from children with viral (n = 10) and bacterial (n = 22) pneumonias was employed (**Figure 1**). Clinical and demographic information are provided in Table 2. These samples were recruited by the PErsonalised Risk assessment in Febrile illness to Optimise Real-life Management across the European Union (PERFORM - https://www.perform2020.org/) and the Diagnosis and Management of Febrile Illness using RNA Personalised Molecular Signature Diagnosis (DIAMONDS - https://www.diamonds2020.eu) consortiums.

All newly generated data from human patients were obtained through harmonized procedures for patient recruitment, classification, clinical data collection, and sample acquisition, processing, and storage across the participating centers. All necessary ethical approvals were obtained from each participating country’s Ethics Committee (EC).

### Method details

#### RNA-seq analysis

For the discovery cohort, RNA was isolated from blood samples collected in PAXgene^TM^ and in Tempus™ RNA tubes. RNA sequencing was conducted on a HiSeq 4000 (Illumina) platform, with library preparation and sequencing of 30 million 75 or 100 bp paired-end reads. The Illumina’s TruSeq RNA Sample Preparation Kit was used for library preparation, and ribosomal and globin RNA depletion was performed using the Illumina® Ribo-Zero Gold kit. For the validation cohort, RNA was isolated using PAXgene blood miRNA isolation kit according to the manufacturer’s instructions (Qiagen). A DNAse treatment was carried out with the RNA clean & concentrator kit (Zymo Research) prior to sequencing. RNA was quantified using RiboGreen (Invitrogen) on the FLUOstar OPTIMA plate reader (BMG Labtech) and the integrity was analyzed on the TapeStation 2200(Agilent, RNA ScreenTape). After a normalization step, a strand specific library preparation was completed using NEBNext® Ultra™ II mRNA kit (NEB) and NEB rRNA/globin depletion probes following manufacturer’s recommendations. Individual libraries were normalized using Qubit, pooled together and diluted. The sequencing was performed using a 150 paired-end configuration in a Novaseq6000 platform.

Quality control of all raw data was carried out using *FastQC* ^55^, alignment and read counting were performed using *STAR* ^56^, alignment filtering was done with *SAMtools* ^57^ and read counting was carried out using *FeatureCounts* ^58^.

#### Differential Expression Analysis

RNAseq data was processed for batch correction using control samples and *COMBAT-Seq* R package ^120^. The *RUVg* method implemented in the *RUVSeq* package of R ^59^ was employed to identified and adjust for other factors of undesirable variation present in the dataset, selecting a *k* = 2 as a number of variables to include. After that, data normalization, differential expression (DE) analysis and covariates correction were carried out using *DESeq* package ^60^. Differentially expressed genes (DEGs) were identified through the Negative Binomial distribution implemented in the *DESeq* package. The fitted model included known covariates such as sex, batch, and type of collection tube as well as the factors of undesirable variation, to correct the differences in gene expression related to these factors. A generalized linear model was fitted, and a *t*-statistic was calculated for each gene. *P*-values were corrected for multiple testing using the Benjamini-Hochberg False Discovery Rate (FDR).

As a validation cohort for the pneumonia *vs*. control study, we downloaded the GSE103119 microarray dataset from the Gene Expression Omnibus (GEO) database. This dataset comprises whole blood gene expression data from children with pneumonia (*n* = 152) as well as healthy controls (*n* = 20). Data were normalized, processed and analyzed using the *limma* R package ^62^. A linear model was fitted, considering the sex as a categorical covariate. Multiple testing correction was performed using the Benjamini-Hochberg False Discovery Rate (FDR).

Principal Component Analysis (PCA) was used to visualize the global transcriptome patterns of both RNA-seq and microarray data. A Spearman test was computed for the correlation indices (ρ) and the associated *P*-values. Wilcoxon test was performed to assess statistical significance between groups.

#### Pathway Analysis

We used the Reactome and GO (Gene Ontology) databases as references to examine biological pathways and processes associated with the DEGs in pneumonia patients. We followed both an over-representation analysis (ORA) and a gene-set enrichment (GSEA) approaches, considering DEGs with a log_2_FC ≥ |1.5| and a adjusted *P*-value < 0.05. The analysis was performed using the *Clusterprofiler* ^63^ R package. To account for multiple test correction, we applied the Benjamini-Hochberg (H–B) procedure, establishing a significant threshold of 0.05.

#### Signature Discovery using LASSO Regression Model

We employed aLeast Absolute Shrinkage and Selection Operator (LASSO) regression model to identify a subset of genes that could serve as a predictive transcriptomic signature differentiating viral from bacterial pneumonia in the RNA-seq cohort. A predictive transcriptomic signature was computed using the R package *glmnet* ^64^. A logistic LASSO regression model was fitted with the alpha parameter set to 1. We selected 36 DEGs filtered based on |log_2_FC| > 1.5, adjusted *P*-value < 0.05, and a Base Mean > 50 as input for the model. To determine the optimal parameter selection for the LASSO regression model, we conducted a 10-fold cross-validation, which helps evaluate the model’s performance and effectively tune the parameters.

To assess the accuracy of the predictive transcriptomic signature, we calculated the area under the receiver operating characteristic curve (AUC) with 95% confidence intervals (CI) using the *pROC* package ^65^ in R. Receiver Operating Characteristic (ROC) curves were generated to graphically represent the model’s true positive rate against the false positive rate. The optimal cut-point value, maximizing sensitivity and specificity, was calculated using the Youden method included in the *OptimalCutPoints* R package ^66^.

#### Signature Validation in an independent cohort

We assessed the performance and accuracy of the 5-transcript signatures using RNAseq data from an additional pediatric cohort (Table 2). Coefficients and intercepts from the LASSO model were applied to the new data to perform ROC analysis and calculate the AUC. We tested different data comparisons: DB *vs*. DV pneumonias, a disaggregation of different DB pneumonias by the causal pathogen (*S.pneumoniae*, *S.aureus* and *S.pyogenes*) *vs*. DV pneumonias and a disaggregation by severity groups based on PICU admission of the patients (mild/moderate *vs.* severe disease).

#### Weighted Gene Correlation Network Analysis (WGCNA)

We conducted a co-expression network analysis using the *WGCNA* package ^67^ to detect clusters of co-expressed genes specifically correlated to viral or bacterial pneumonia. We selected all patients with DV (*n* = 9) and DB (*n* = 40) pneumonia for the analysis. Only protein-coding genes exhibiting the highest expression variance among samples (the top 25%) were included, totaling 4,831 genes. Normalized gene expression data were used to construct a signed weighted correlation network. A matrix of correlations between all pairs of selected genes was generated from the expression values and converted into an adjacency matrix with a power function. We determined a soft-thresholding power of 20 (maximum model fitting index), selected on the criterion of scale-free topology after evaluating several candidate powers (**Figure S5**).

We calculated the Topological Overlap Matrix (TOM) and the corresponding dissimilarity (1–TOM) values. For module detection and merging, we showed a minimum module size of 30, and a cut height threshold of 20, respectively. Correlation between module eigengenes and the viral/bacterial pneumonia was calculated to identify modules of co-expressed genes significantly associated with the phenotype (gene significance, or GS) and using the viral category as reference. Module Membership (MM) measured intramodular connectivity, and the most interconnected gene (hub gene) named the modules.

The biological pathways represented by each of the significantly correlated modules were investigated through an over-representation analysis with the *Clusterprofiler* R package ^63^, with terms from GO and Reactome databases as references.

All graphics were created using R software v.4.2.0 (www-r-project.org).

## Supporting information

Key Resources Table

## Data Availability

All data produced in the present study are available upon reasonable request to the authors

## Data and Code Availability

RNA-seq data supporting the findings of the present study are available under request.

## Acknowledgements

The authors would like to express their appreciation to the study investigators of GENDRES network (www.gendres.org) (Annex), as well as the nursery and laboratory service at the Hospital Clínico Universitario de Santiago de Compostela, for their invaluable dedication and support. This research project was made possible through the access granted by the Galician Supercomputing Center (CESGA) to its supercomputing infrastructure. The supercomputer FinisTerrae III and its permanent data storage system have been funded by the Spanish Ministry of Science and Innovation, the Galician Government, and the European Regional Development Fund (ERDF). This work was supported by the European Seventh Framework Programme for Research and Technological Development (FP7) under EUCLIDS project (Grant Agreement number 279185) and the European Union’s Horizon 2020 research and innovation programme under Grant Agreement Nos. 668303 (PERFORM) and 848196 (DIAMONDS). This study also received support by i) ISCIII: TRINEO: PI22/00162; DIAVIR: DTS19/00049; Resvi-Omics: PI19/01039 (to A.S.), ReSVinext: PI16/01569, Enterogen: PI19/01090, OMI-COVI-VAC: PI22/00406 (to F.M.-T.), cofinanciados FEDER, ii) GAIN: IN607B 2020/08 and IN607A 2023/02 (to A.S.), GEN-COVID (IN845D 2020/23 (to F.M.-T.), IIN607A2021/05 (to F.M.-T.); iii) ACIS: BI-BACVIR (PRIS-3, to A.S.), CovidPhy (SA 304 C, to A.S.); iv) Spanish Ministry of Science and Innovation (MCIN)/Spanish Research Agency (AEI) (PID2022-142156OB-I00, to AG-C); and v) consorcio Centro de Investigación Biomédica en Red de Enfermedades Respiratorias (CB21/06/00103; to A.S. and F.M.-T.). AGC is supported by the Miguel Servet programme (CP23/00080) contract, funded by the Instituto de Salud Carlos III (ISCIII) and co-funded by the European Union. The funders were not involved in the study design, collection, analysis, interpretation of data, the writing of this article or the decision to submit it for publication.

## GENDRES consortium

**GENDRES is a network of hospital and primary care centers created in 2010 for the study of acute respiratory infections. It is primarily focused on the omics approach to these infections to understand their pathogenesis, genetic susceptibility factors, prognosis, and the detection of diagnostic or predictive biomarkers. This is achieved through the collection of samples and clinical data from pediatric subjects presenting with acute respiratory symptoms in participating centers.**

**Researchers of GENDRES network**

Miriam Cebey López, Antonio Salas Ellacuriaga, Ana Vega Gliemmo, José Peña Guitián, Alexa Regueiro, Antonio Justicia Grande, Leticia Pías Peleteiro, María López Sousa, María Jose de Castro, Carmen Curros Novo, Elena Rodrigo, Miriam Puente Puig, Rosaura Leis Trabazo, Nazareth Martinón Torres, Alberto Gómez Carballa, Jacobo Pardo Seco, Sara Pischedda, José María Martinón Sánchez, Belén Mosquera Pérez, Isabel Villanueva González, Lorenzo Redondo Collazo, Carmen Rodríguez-Tenreiro, María del Sol Porto Silva and Federico Martinón Torres (Área Asistencial Integrada de Pediatría and GENVIP, Hospital Clínico Universitario, Santiago de Compostela); Máximo Francisco Fraga Rodríguez, Orlando Fernández Lago, José Ramón Antúnez (Biobank, Servicio Anatomía Patológica, Hospital Clínico Universitario, Santiago de Compostela); Enrique Bernaola Iturbe, Laura Moreno Galarraga, Jorge Álvarez, Mercedes Herranz, Francisco Gil, Eva Gembero, Jorge Rodríguez (Hospital Materno Infantil Virgen del Camino, Pamplona); Teresa González López, Delfina Suarez Vázquez, Ángela Vázquez Vázquez, Susana Rey García, Nathalie Carreira Sande, Ana López Fernández, Nuria Romero Pérez (Complejo Hospitalario Universitario de Orense); José Antonio Couceiro Gianzo, Nazareth Fuentes Perez (Complejo Hospitalario Universitario de Pontevedra); Francisco Giménez Sánchez, Miguel Sánchez Forte (Hospital Torrecárdenas, Almería); Cristina Calvo Rey, María Luz García García, Iciar Olabarrieta Arnal, Adelaida Fernández Rincón (Hospital Severo Ochoa de Madrid); Ignacio Oulego Erroz, David Naranjo Vivas, Santiago Lapeña, Paula Alonso Quintela, Jorge Martínez Sáenz de Jubera, Estibaliz Garrido García (Hospital de León); Ana Grande Tejada (Hospital Materno Infantil de Badajoz); Cristina Calvo Monge, Eider Oñate Vergara (Hospital de Donostia, San Sebastián); Jesús de la Cruz Moreno, Mª Carmen Martínez Padilla, Beatriz Jiménez Jurado, Carmen Santiago Gutiérrez, María Esther Vidaurreta del Castillo (Complejo Hospitalario de Jaén); Manuel Baca Cots (Hospital Quirón, Málaga), David Moreno Pérez, Ana Cordón Martínez, Antonio Urda Cardona, José Miguel Ramos Fernández, Esmeralda Núñez Cuadros (Hospital Carlos Haya, Málaga); Susana Beatriz Reyes, María Cruz León León, Santiago Alfayate (Hospital Virgen de la Arrixaca, Murcia); Cristina Calvo, Carlos Grasa, Cristian Quintana Ortega, Leticia La Banda Montalvo, María Lopez Cerdán, Ana Dominguez Castells (Hospital Universitario La Paz, Madrid); Francisco Giménez Sánchez (Hospital Inmaculada); Andrés J. Alcaraz Romero, Diego Bautista Lozano, Sara Uillen Martin (Hospital Universitario de Getafe); Roi Piñeiro (Hospital General de Villalba); Juan Ignacio Sánchez Díaz, Alba Palacios Cuesta (Hospital 12 Octubre); Elvira González Salas, Sira Fernández De Miguel (Hospital Clínico de Salamanca); Belen Joyanes Abancens, Esther Aleo Lujan (Hospital clínico San Carlos); Alfredo Tagarro García, María Luisa Herreros, Rut del Valle, Libertad Latorre Navarro (Hospital Universitario Infanta Sofía); María Concepción Zazo Sanchidrián, Mariano Esteban, Marta González Lorenzo, Mª Carmen Vicent Castello (Hospital General Universitario de Alicante); Lorena Moreno Requena, Juan Luis Santos Pérez (Complejo Hospitalario Universitario de Granada); César Gavilán Martín, Lucía González-Moro Azorín (Hospital Universitario de San Juan de Alicante); Monterrat López Franco, Manuel Silveira Cancela (Hospital de Burela); María José Cilleruelo Ortega, Luz Golmayo, Francisca Portero Azorín (Hospital Universitario Puerta de Hierro-Majadahonda); Andrés Concha Torre, Lucía Rodríguez García (Hospital Universitario Central de Asturias); Carlos Rodrigo Gonzalo de Liria, Andrés Antón Pagarolas (Hospital Vall d’Hebron); Maria Méndez, Cristina Prat (Hospital Germans Trias i Pujol); Jesús López-Herce, Miriam García Samprudencio, Gema Manrique Martín, Paula García Casas, Débora Sanz Álvarez (Hospital General Universitario Gregorio Marañón); María Jesús Cabero (Hospital Valdecilla); Miguel Lillo Lillo, Marta Pareja (Hospital General de Albacete); Pablo Rojo, Cristina Epalza (Hospital 12 Octubre); Adriana Navas Carretero, Estefanía Barral, Miriam Herrera (Hospital Infanta Leonor); Elvira Cobo Vázquez (Hospital Universitario Fundación de Alcorcón); Elena del Castillo Navío (Hospital Materno Infantil de Badajoz); Patricia Flores Perez (Hospital del Niño Jesús); Paula García Casas (Hospital Ramón y Cajal); Esteban Gómez Sánchez, Juan Valencia Ramos (Hospital Universitario de Burgos); Francisco Javier Pilar Orive, Elva Rodríguez Merino (Hospital Universitario Cruces); Ana Pérez Aragón (Hospital Universitario Virgen de las Nieves de Granada); Mª Yolanda Ruiz del Prado (Hospital San Pedro); David Moreno, Beatriz Carazo (Hospital Carlos Haya); Jordi Antón (Hospital Sant Joan de Déu); María teresa Rives Ferreiro (Hospital Universitario de Navarra). Further details may be consulted at http://www.gendres.org, and www.genvip.eu.

## EUCLIDS consortium

**Imperial College partner (UK)**

Members of the EUCLIDS Consortium at Imperial College London (UK)

<u>Principal and co-investigators</u>

Michael Levin (grant application, EUCLIDS Coordinator)

Dr. Lachlan Coin (bioinformatics)

Stuart Gormley (clinical coordination)

Shea Hamilton (proteomics)

Jethro Herberg (grant application, PI)

Bernardo Hourmat (project management)

Clive Hoggart (statistical genomics)

Myrsini Kaforou (bioinformatics)

Vanessa Sancho-Shimizu (genetics)

Victoria Wright (grant application, scientific coordination)

<u>Consortium members at Imperial College</u>

Amina Abdulla

Paul Agapow

Maeve Bartlett

Evangelos Bellos

Hariklia Eleftherohorinou

Rachel Galassini

David Inwald

Meg Mashbat

Stefanie Menikou

Sobia Mustafa

Simon Nadel

Rahmeen Rahman

Clare Thakker

<u>EUCLIDS UK Clinical Network</u>

Poole Hospital NHS Foundation Trust, Poole: Dr S Bokhandi (PI), Sue Power, Heather Barham

Cambridge University Hospitals NHS Trust, Cambridge: Dr N Pathan (PI), Jenna Ridout, Deborah White, Sarah Thurston

University Hospital Southampton, Southampton: Prof S Faust (PI), Dr S Patel (co-investigator), Jenni McCorkell.

Nottingham University Hospital NHS Trust: Dr P Davies (PI), Lindsey Crate, Helen Navarra, Stephanie Carter

University Hospitals of Leicester NHS Trust, Leicester: Dr R Ramaiah (PI), Rekha Patel

Portsmouth Hospitals NHS Trust, London: Dr Catherine Tuffrey (PI), Andrew Gribbin, Sharon McCready

Great Ormond Street Hospital, London: Dr Mark Peters (PI), Katie Hardy, Fran Standing, Lauran O’Neill, Eugenia Abelake

King’s College Hospital NHS Foundation Trust, London; Dr Akash Deep (PI), Eniola Nsirim

Oxford University Hospitals NHS Foundation Trust, Oxford Prof A Pollard (PI), Louise Willis, Zoe Young

Kettering General Hospital NHS Foundation Trust, Kettering: Dr C Royad (PI), Sonia White

Central Manchester NHS Trust, Manchester: Dr PM Fortune (PI), Phil Hudnott

**SERGAS Partner (Spain)**

<u>Principal Investigators</u>

Federico Martinón-Torres^1^

Antonio Salas^1,2^

<u>GENVIP/GenPoB RESEARCH GROUPS (in alphabetical order):</u>

Fernando Álvez González^1^, Ruth Barral-Arca^1,2^, Miriam Cebey-López^1^, María José Curras-Tuala^1,2^, Natalia García^1^, Luisa García Vicente^1^, Alberto Gómez-Carballa^1,2^, Jose Gómez Rial^1^, Andrea Grela Beiroa^1^, Antonio Justicia Grande^1^, Pilar Leboráns Iglesias^1^, Alba Elena Martínez Santos^1^, Federico Martinón-Torres^1^, Nazareth Martinón-Torres^1^, José María Martinón Sánchez^1^, Beatriz Morillo Gutiérrez^1^, Belén Mosquera Pérez^1^, Pablo Obando Pacheco^1^, Jacobo Pardo-Seco^1,2^, Sara Pischedda^1,2^, Irene Rivero Calle^1^, Carmen Rodríguez-Tenreiro^1^, Lorenzo Redondo-Collazo^1^, Antonio Salas Ellacuriaga^1,2^, Sonia Serén Fernández^1^, María del Sol Porto Silva^1^, Ana Vega^1,3^, Jose Manuel Fernández García^3^, María Elena Gamborino Caramés^3^, María Sol Rodríguez Calvo^3^, Marta Aldonza Torres^3^, Vanesa Álvarez Iglesias^3^, Carmen Curros Novo^3^, Isabel Ferreirós Vidal^3^, Narmeen Mallah^3^, Laura Navarro Ramón^3^, Isabel Rego Lijo^3^, Alba Camino Mera^3^, Lúa Castelo Martínez^3^, Ana Isabel Dacosta Urbietav, Wiktor Dominik Nowak^3^, Julia García Currás^3^, Nour El Zaharaa Mallah^3^, Julián Montoto Louzao^3^, Sara Rey Vázquez^3^, Sandra Viz Lasheras^3^, Patricia Regueiro Casuso^3^, Miriam Taboada Puga^3^, Lucía Vilanova Trillo^1^

1. Translational Pediatrics and Infectious Diseases, Pediatrics Department, Hospital Clínico Universitario de Santiago, Santiago de Compostela, Spain, and GENVIP Research Group (www.genvip.org), Instituto de Investigación Sanitaria de Santiago, Galicia, Spain.
2. Unidade de Xenética, Departamento de Anatomía Patolóxica e Ciencias Forenses, Instituto de Ciencias Forenses, Facultade de Medicina, Universidade de Santiago de Compostela, and Genética de Poblaciones en Biomedician (GenPop) Research Group, Instituto de Investigaciones Sanitarias (IDIS), Hospital Clínico Universitario de Santiago, Galicia, Spain
3. Fundación Pública Galega de Medicina Xenómica, Servizo Galego de Saúde (SERGAS), Instituto de Investigaciones Sanitarias (IDIS), and Grupo de Medicina Xenómica, Centro de Investigación Biomédica en Red de Enfermedades Raras (CIBERER), Universidade de Santiago de Compostela (USC), Santiago de Compostela, Spain

<u>EUCLIDS SPANISH CLINICAL NETWORK:</u>

Susana Beatriz Reyes^1^, María Cruz León León^1^, Álvaro Navarro Mingorance^1^, Xavier Gabaldó Barrios^1^, Eider Oñate Vergara^2^, Andrés Concha Torre^3^, Ana Vivanco^3^, Reyes Fernández^3^, Francisco Giménez Sánchez^4^, Miguel Sánchez Forte^4^, Pablo Rojo^5^, J.Ruiz Contreras^5^, Alba Palacios^5^, Cristina Epalza Ibarrondo^5^, Elizabeth Fernandez Cooke^5^, Marisa Navarro^6^, Cristina Álvarez Álvarez^6^, María José Lozano^6^, Eduardo Carreras^7^, Sonia Brió Sanagustín^7^, Olaf Neth^8^, Mª del Carmen Martínez Padilla^9^, Luis Manuel Prieto Tato^10^, Sara Guillén^10^, Laura Fernández Silveira^11^, David Moreno^12^.

1. Hospital Clínico Universitario Virgen de la Arrixaca; Murcia, Spain.
2. Hospital de Donostia; San Sebastián, Spain.
3. Hospital Universitario Central de Asturias; Asturias, Spain.
4. Complejo Hospitalario Torrecárdenas; Almería, Spain.
5. Hospital Universitario 12 de Octubre; Madrid, Spain.
6. Hospital General Universitario Gregorio Marañón; Madrid, Spain.
7. Hospital de la Santa Creu i Sant Pau; Barcelona, Spain.
8. Hospital Universitario Virgen del Rocío; Sevilla, Spain.
9. Complejo Hospitalario de Jaén; Jaén, Spain.
10. Hospital Universitario de Getafe; Madrid, Spain.
11. Hospital Universitario y Politécnico de La Fe; Valencia, Spain.
12. Hospital Regional Universitario Carlos Haya; Málaga, Spain.

**Members of the Pediatric Dutch Bacterial Infection Genetics (PeD-BIG) network (the Netherlands)**

<u>Steering committee:</u>

Coordination: R. de Groot ^1^, A.M. Tutu van Furth ^2^, M. van der Flier ^1^

Coordination Intensive Care: N.P. Boeddha ^3^, G.J.A. Driessen ^3^, M. Emonts ^3^, J.A. Hazelzet ^3^

Other members: T.W. Kuijpers ^5^, D. Pajkrt ^5^, E.A.M. Sanders ^4^, D. van de Beek ^6^, A. van der Ende^6^

Trial coordinator: H.L.A. Philipsen ^1^

Local investigators (in alphabetical order)

A.O.A. Adeel ^7^, M.A. Breukels ^8^, D.M.C. Brinkman ^9^, C.C.M.M. de Korte ^10^, E. de Vries ^11^, W.J. de Waal ^12^, R. Dekkers ^13^, A. Dings-Lammertink ^14^, R.A. Doedens ^15^, A.E. Donker ^16^, M. Dousma^17^, T.E. Faber ^18^, G.P.J.M. Gerrits^19^, J.A.M. Gerver ^20^, J. Heidema ^21^, J. Homan-van der Veen ^22^, M.A.M. Jacobs ^23^, N.J.G. Jansen ^4^, P. Kawczynski ^24^, K. Klucovska ^25^, M.C.J. Kneyber ^26^, Y. Koopman-Keemink ^27^, V.J. Langenhorst ^28^, J. Leusink ^29^, B.F. Loza ^30^, I.T. Merth ^31^, C.J. Miedema ^32^, C. Neeleman ^1^, J.G. Noordzij ^33^, C.C. Obihara ^34^, A.L.T. van Overbeek – van Gils ^35^, G.H. Poortman ^36^,S.T. Potgieter ^37^, J. Potjewijd ^38^, P.P.R. Rosias ^39^, T. Sprong ^19^, G.W. ten Tussher ^40^, B.J. Thio ^41^, G.A. Tramper-Stranders ^42^, M. van Deuren ^1^, H. van der Meer ^2^, A.J.M. van Kuppevelt ^43^, A.M. van Wermeskerken ^44^, W.A. Verwijs ^45^, T.F.W. Wolfs ^4^.

1. Radboud University Medical Center – Amalia Children’s Hospital, Nijmegen, The Netherlands
2. Vrije Universiteit University Medical Center, Amsterdam, The Netherlands
3. Erasmus Medical Center – Sophia Children’s Hospital, Rotterdam, The Netherlands
4. University Medical Center Utrecht – Wilhelmina Children’s Hospital, Utrecht, The Netherlands
5. Academic Medical Center – Emma Children’s Hospital, University of Amsterdam, Amsterdam, The Netherlands
6. Academic Medical Center, University of Amsterdam, Amsterdam, The Netherlands
7. Kennemer Gasthuis, Haarlem, The Netherlands
8. Elkerliek Hospital, Helmond, The Netherlands
9. Alrijne Hospital, Leiderdorp, The Netherlands
10. Beatrix Hospital, Gorinchem, The Netherlands
11. Jeroen Bosch Hospital, ‘s-Hertogenbosch, The Netherlands
12. Diakonessenhuis, Utrecht, The Netherlands
13. Maasziekenhuis Pantein, Boxmeer, The Netherlands
14. Gelre Hospitals, Zutphen, The Netherlands
15. Martini Hospital, Groningen, The Netherlands
16. Maxima Medical Center, Veldhoven, The Netherlands
17. Gemini Hospital, Den Helder, The Netherlands
18. Medical Center Leeuwarden, Leeuwarden, The Netherlands
19. Canisius-Wilhelmina Hospital, Nijmegen, The Netherlands
20. Rode Kruis Hospital, Beverwijk, The Netherlands
21. St. Antonius Hospital, Nieuwegein, The Netherlands
22. Deventer Hospital, Deventer, The Netherlands
23. Slingeland Hospital, Doetinchem, The Netherlands
24. Refaja Hospital, Stadskanaal, The Netherlands
25. Bethesda Hospital, Hoogeveen, The Netherlands
26. University Medical Center Groningen, Beatrix Children’s hospital, Groningen, The Netherlands
27. Haga Hospital – Juliana Children’s Hospital, Den Haag, The Netherlands
28. Isala Hospital, Zwolle, The Netherlands
29. Bernhoven Hospital, Uden, The Netherlands
30. VieCuri Medical Center, Venlo, The Netherlands
31. Ziekenhuisgroep Twente, Almelo-Hengelo, The Netherlands
32. Catharina Hospital, Eindhoven, The Netherlands
33. Reinier de Graaf Gasthuis, Delft, The Netherlands
34. ETZ Elisabeth, Tilburg, The Netherlands
35. Scheper Hospital, Emmen, The Netherlands
36. St. Jansdal Hospital, Hardewijk, The Netherlands
37. Laurentius Hospital, Roermond, The Netherlands
38. Isala Diaconessenhuis, Meppel, The Netherlands
39. Zuyderland Medical Center, Sittard-Geleen, The Netherlands
40. Westfriesgasthuis, Hoorn, The Netherlands
41. Medisch Spectrum Twente, Enschede, The Netherlands
42. St. Franciscus Gasthuis, Rotterdam, The Netherlands
43. Streekziekenhuis Koningin Beatrix, Winterswijk, The Netherlands
44. Flevo Hospital, Almere, The Netherlands
45. Zuwe Hofpoort Hospital, Woerden, The Netherlands

**Swiss Pediatric Sepsis Study**

<u>Steering Committee</u>

Luregn J Schlapbach, MD, FCICM^1,2,3^, Philipp Agyeman, MD^1^, Christoph Aebi, MD^1^, Christoph Berger, MD^1^, Luregn J Schlapbach, MD, FCICM^1,2,3^, Philipp Agyeman, MD^1^, Christoph Aebi, MD^1^, Eric Giannoni, MD^4,5^, Martin Stocker, MD^6^, Klara M Posfay-Barbe, MD^7^, Ulrich Heininger, MD^8^, Sara Bernhard-Stirnemann, MD^9^, Loher, MD^10^, Christian Kahlert, MD^10^, Paul Hasters, MD^11^, Christa Relly, MD^12^, Walter Baer, MD^13^, Christoph Berger, MD^12^ for the Swiss Pediatric Sepsis Study

1. Department of Pediatrics, Inselspital, Bern University Hospital, University of Bern, Switzerland
2. Paediatric Critical Care Research Group, Mater Research Institute, University of Queensland, Brisbane, Australia
3. Paediatric Intensive Care Unit, Lady Cilento Children’s Hospital, Children’s Health Queensland, Brisbane, Australia
4. Service of Neonatology, Lausanne University Hospital, Lausanne, Switzerland
5. Infectious Diseases Service, Lausanne University Hospital, Lausanne, Switzerland
6. Department of Pediatrics, Children’s Hospital Lucerne, Lucerne, Switzerland
7. Pediatric Infectious Diseases Unit, Children’s Hospital of Geneva, University Hospitals of Geneva, Geneva, Switzerland
8. Infectious Diseases and Vaccinology, University of Basel Children’s Hospital, Basel, Switzerland
9. Children’s Hospital Aarau, Aarau, Switzerland
10. Division of Infectious Diseases and Hospital Epidemiology, Children’s Hospital of Eastern Switzerland St. Gallen, St. Gallen, Switzerland
11. Department of Neonatology, University Hospital Zurich, Zurich, Switzerland
12. Division of Infectious Diseases and Hospital Epidemiology, and Children’s Research Center, University Children’s Hospital Zurich, Switzerland
13. Children’s Hospital Chur, Chur, Switzerland

**Liverpool Partner**

<u>Principal Investigators</u>

Enitan Carrol^1^

Stéphane Paulus ^2^

EUCLIDS research team (in alphabetical order):

Hannah Frederick^3^, Rebecca Jennings^3^, Joanne Johnston^3^, Rhian Kenwright^3^

1. Department of Clinical Infection, Microbiology and Immunology, University of Liverpool Institute of Infection, Veterinary and Ecological Sciences, Liverpool, England
2. Alder Hey Children’s Hospital, Department of Infectious Diseases, Eaton Road, Liverpool, L12 2AP
3. Alder Hey Children’s Hospital, Clinical Research Business Unit, Eaton Road, Liverpool, L12 2AP

**Micropathology Ltd**

Colin G Fink^1,2^

Elli Pinnock^1^

1. Micropathology Ltd Research and Diagnosis
2. University of Warwick

**Newcastle partner**

<u>Principle Investigator</u>

Marieke Emonts^1,2^

<u>Co-Investigator</u>

Rachel Agbeko^1,3^

1. Institute of Cellular Medicine, Newcastle University, Newcastle upon Tyne, United Kingdom
2. Paediatric Infectious Diseases and Immunology Department, Newcastle upon Tyne Hospitals Foundation Trust, Great North Children’s Hospital, Newcastle upon Tyne, United Kingdom
3. Paediatric Intensive Care Unit, Newcastle upon Tyne Hospitals Foundation Trust, Great North Children’s Hospital, Newcastle upon Tyne, United Kingdom

**Gambian partner**

<u>Principal Investigator and West African study oversight:</u>

Suzanne Anderson^1^

<u>Clinical research fellow and study co-ordinator</u>

Fatou Secka^1^

<u>Additional Gambia site team (consortium members):</u>

Kalifa Bojang^1^: co-PI

Isatou Sarr^1^: Senior laboratory technician

Ngane Kebbeh^1^: Junior laboratory technician

Gibbi Sey^1^: lead research nurse Medical Research Council Clinic

Momodou Saidykhan^1^: lead research nurse Edward Francis Small Teaching Hospital

Fatoumatta Cole^1^: Data manager

Gilleh Thomas^1^: Data manager

Martin Antonio^1^: Local collaborator

1. Medical Research Council Unit Gambia, PO Box 27, Banjul, The Gambia

**Austrian partner**

<u>Principal investigator</u>

Werner Zenz^1^

<u>Co-Investigators</u>

Daniela S. Klobassa^1^, Alexander Binder^1^, Nina A. Schweintzger^1^, Manfred Sagmeister^1^

1. University Clinic of Paediatrics and Adolescent Medicine, Department of General Paediatrics, Medical University Graz, Austria

Austrian network, participating centres in Austria, Germany, Italy, Serbia, Lithuania, patient recruitment (in alphabetical order):

Hinrich Baumgart^1^, Markus Baumgartner^2^, Uta Behrends^3^, Ariane Biebl^4^, Robert Birnbacher^5^, Jan-Gerd Blanke^6^, Carsten Boelke^7^, Kai Breuling^3^, Jürgen Brunner^8^, Maria Buller^9^, Peter Dahlem^10^, Beate Dietrich^11^, Ernst Eber^12^, Johannes Elias^13^, Josef Emhofer^2^, Rosa Etschmaier^14^, Sebastian Farr^15^, Ylenia Girtler^16^, Irina Grigorow^17^, Konrad Heimann^18^, Ulrike Ihm^19^, Zdenek Jaros^20^, Hermann Kalhoff^21^, Wilhelm Kaulfersch^22^, Christoph Kemen^23^, Nina Klocker^24^, Bernhard Köster^25^, Benno Kohlmaier^26^, Eleni Komini^27^, Lydia Kramer^3^, Antje Neubert^28^, Daniel Ortner^29^, Lydia Pescollderungg^16^, Klaus Pfurtscheller^30^, Karl Reiter^31^, Goran Ristic^32^, Siegfried Rödl^30^, Andrea Sellner^26^, Astrid Sonnleitner^26^, Matthias Sperl^33^, Wolfgang Stelzl^34^, Holger Till^1^, Andreas Trobisch^26^, Anne Vierzig^35^, Ulrich Vogel^12^, Christina Weingarten^36^, Stefanie Welke^37^, Andreas Wimmer^38^, Uwe Wintergerst^39^, Daniel Wüller^40^, Andrew Zaunschirm^41^, Ieva Ziuraite^42^, Veslava Žukovskaja^42^

1. Department of Pediatric and Adolescence Surgery, Division of General Pediatric Surgery, Medical University Graz, Austria
2. Department of Pediatrics, General Hospital of Steyr, Austria
3. Department of Pediatrics/Department of Pediatric Surgery, Technische Universität München (TUM), Munich, Germany
4. Department of Pediatrics, Kepler University Clinic, Medical Faculty of the Johannes Kepler University, Linz, Austria
5. Department of Pediatrics and Adolesecent Medicine LKH Villach, Austria
6. Department of Pediatrics and Adolescent Medicine and Neonatology, Hospital Ludmillenstift, Meppen, Germany
7. Hospital for Children’s and Youth Medicine, Oberschwabenklinik, Ravensburg, Germany
8. Department of Pediatrics, Medical University Innsbruck, Austria
9. Clinic for Paediatrics and Adolescents Medicine, Sana Hanse-Klinikum Wismar, Germany
10. Departement of Pediatrics, Medical Center Coburg, Germany
11. University Medicine Rostock, Department of Pediatrics (UKJ), Rostock, Germany
12. Department of Pulmonology, Medical University Graz, Austria
13. Institute for Hygiene and Microbiology, University of Würzburg, Germany
14. Clinical Institute of Medical and Chemical Laboratory Diagnostics, Medical University Graz, Austria
15. Department of Pediatric Orthopedics and Adult Foot and Ankle Surgery, Orthopedic Hospital Speising, Vienna, Austria
16. Department of Paediatrics, Regional Hospital Bolzano, Italy
17. Department of Pediatrics and Adolescent Medicine, General Hospital Hochsteiermark/Leoben, Austria
18. Department of Neonatology and Paediatric Intensive Care, Children’s University Hospital, RWTH Aachen, Germany
19. Paediatric Intensive Care Unit, Department of Paediatric Surgery, Donauspital Vienna, Austria
20. Department of Pediatrics, General Public Hospital, Zwettl, Austria
21. Pediatric Clinic Dortmund, Germany
22. Department of Pediatrics and Adolescent Medicine, Klinikum Klagenfurt am Wörthersee, Klagenfurt, Austria
23. Catholic Children’s Hospital Wilhelmstift, Department of Pediatrics, Hamburg, Germany
24. Department of Pediatrics, Krankenhaus Dornbirn, Austria
25. Children’s Hospital Luedenscheid, Maerkische Kliniken, Luedenscheid, Germany
26. Department of General Paediatrics, Medical University Graz, Austria
27. Department of Paediatrics, Schwarzwald-Baar-Hospital, Villingen-Schwenningen, Germany
28. Department of Paediatrics and Adolescents Medicine, University Hospital Erlangen, Germany
29. Department of Pediatrics and Adolescent Medicine, Medical University of Salzburg, Austria
30. Paediatric Intensive Care Unit, Medical University Graz, Austria
31. Dr. von Hauner Children’s Hospital, Ludwig-Maximilians-Universitaet, Munich, Germany
32. Mother and Child Health Care Institute of Serbia, Belgrade, Serbia
33. Department of Pediatric and Adolescence Surgery, Division of Pediatric Orthopedics, Medical University Graz, Austria
34. Department of Pediatrics, Academic Teaching Hospital, Landeskrankenhaus Feldkirch, Austria
35. University Children’s Hospital, University of Cologne, Germany
36. Department of Pediatrics and Adolescent Medicine Wilheminenspital, Vienna, Austria
37. Department of Pediatric Surgery, Municipal Hospital Karlsruhe, Germany
38. Hospital of the Sisters of Mercy Ried, Department of Pediatrics and Adolescent Medicine, Ried, Austria
39. Hospital St. Josef, Braunau, Austria
40. Christophorus Kliniken Coesfeld Clinic for Pediatrics, Coesfeld, Germany
41. Department of Paediatrics, University Hospital Krems, Karl Landsteiner University of Health Sciences, Krems, Austria
42. Children‘s Hospital, Affiliate of Vilnius University Hospital Santariskiu Klinikos, Lithuania

**DIAMONDS Consortium**

https://www.diamonds2020.eu/

**Imperial College (Coordinating Centre) (UK)**

Chief investigator/DIAMONDS coordinator:

Michael Levin^1^

Principal and co-investigators (alphabetical order)^1^

Aubrey Cunnington; Jethro Herberg; Myrsini Kaforou; Victoria Wright

Section of Paediatric Infectious Diseases Research Group (alphabetical order)^1^ Evangelos Bellos; Claire Broderick; Samuel Channon-Wells; Samantha Cooray; Tisham De (database work package lead); Giselle D’Souza; Leire Estramiana Elorrieta; Diego Estrada-Rivadeneyra; Rachel Galassini (Clinical Trial Manager); Dominic Habgood-Coote; Shea Hamilton (Proteomics); Heather Jackson; James Kavanagh; Mahdi Moradi Marjaneh; Stephanie Menikou; Samuel Nichols; Ruud Nijman; Harsita Patel; Ivana Pennisi; Oliver Powell; Ruth Reid; Priyen Shah; Ortensia Vito; Elizabeth Whittaker; Clare Wilson; Rebecca Womersley

Recruitment team at Imperial College Healthcare NHS Trust, London (alphabetical order)^2^

Amina Abdulla; Sarah Darnell; Sobia Mustafa

Engineering Team

Pantelis Georgiou^3^ (engineering lead); Jesus-Rodriguez Manzano^4^; Nicolas Moser^3^; Ivana Pennisi^1^

1. Section of Paediatric Infectious Disease, Imperial College London, Norfolk Place, London W2 1PG, UK
2. Children’s Clinical Research Unit, St Mary’s Hospital, Praed Street, London W2 1NY, UK
3. Imperial College London, Department of Electrical and Electronic Engineering, South Kensington Campus, London, SW7 2AZ, UK
4. Imperial College London, Department of Infectious Disease, Section of Adult Infectious Disease, Hammersmith Campus, London, W12 0NN, UK

**UK Non-Consortium Clinical Recruiting Sites**

<u>Evelina London Children’s Hospital, Guy’s and St Thomas’ NHS Foundation Trust; King’s College London [combined]</u>

Michael Carter^1,2^ (principal investigator); Shane Tibby^1,2^ (co-investigator)

Recruitment team (alphabetical order): Jonathan Cohen^1^; Francesca Davis^1^; Julia Kenny^1^; Paul Wellman^1^; Marie White^1^

Laboratory team (alphabetical order): Matthew Fish^3^; Aislinn Jennings^4^; Manu Shankar-Hari^3,4^

1. Evelina London Children’s Hospital, Guy’s and St Thomas’ NHS Foundation Trust, London, UK
2. Department of Women and Children’s Health, School of Life Course Sciences, King’s College London, UK
3. Department of Infectious Diseases, School of Immunology and Microbial Sciences, King’s College London, London, UK
4. Department of Intensive Care Medicine, Guy’s and St Thomas’ NHS Foundation Trust, London, UK

<u>University Hospitals Sussex</u>

Katy Fidler^1^ (principal investigator); Dan Agranoff^2^ (co-investigator) Recruitment team; Vivien Richmond^1,3^, Mathhew Seal^2^

1. Royal Alexandra Children’s Hospital, University Hospitals Sussex, Brighton, UK
2. Dept of Infectious Diseases, University Hospitals Sussex, Brighton, UK
3. Research Nurse team, University Hospitals Sussex, Brighton, UK

<u>University Hospital Southampton NHS Foundation Trust</u>

Saul Faust^1^ (principal investigator); Dan Owen^1^ (co-investigator);

Recruitment team; Ruth Ensom^2^; Sarah McKay^2^; Diana Mondo^3^, Mariya Shaji^3^; Rachel Schranz^3^ (alphabetical order)

1. NIHR Southampton Clinical Research Facility, University Hospital Southampton NHS Foundation Trust and University of Southampton, UK
2. NIHR Southampton Clinical Research Facility, University Hospital Southampton NHS Foundation Trust, UK
3. Department of R&D, University Hospital Southampton NHS Foundation Trust, UK

<u>Barts Health NHS Trust</u>

Prita Rughnani^1, 2, 3^ (principal investigator 2020-2021); Amutha Anpananthar^1, 2, 3^ (principal investigator 2021-to date); Susan Liebeschuetz^2^ (co-investigator), Anna Riddell^1^ (co-investigator)

Recruitment team; Nosheen Khalid^1, 3^,Ivone Lancoma Malcolm, Teresa Simagan^3^ (alphabetical order)

1. Royal London Hospital, Whitechapel Rd, London E1 1FR, UK
2. Newham University Hospital, Glen Rd, London E13 8SL, UK
3. Whipps Cross University Hospital, Whipps Cross Road, London, E11 1NR, UK

<u>Great Ormond Street Hospital for Children NHS Foundation Trust</u>

Mark Peters^1,2^ (principal investigator); Alasdair Bamford^1,2^ (co-investigator) Recruitment team; Lauran O’Neill^1^

1. Great Ormond Street Hospital, London, WC1N 3JH, UK
2. UCL Great Ormond St Institute of Child Health, WC1N 1EH, UK

<u>Cambridge University Hospitals NHS Foundation Trust</u>

Nazima Pathan^1,2^ (principal investigator)

Recruitment team; Esther Daubney^1^, Deborah White^1^ (alphabetical order)

1. Addenbrooke’s Hospital, Hills Road, Cambridge CB2 0QQ, UK
2. Department of Paediatrics, University of Cambridge, Cambridge CB2 0QQ, UK

<u>University College London Hospitals NHS Foundation Trust</u>

Melissa Heightman^1^ (principal investigator); Sarah Eisen^1^ (co-investigator)

Recruitment team; Terry Segal^1^, Lucy Wellings^1^ (alphabetical order)

1. University College London Hospital, Euston Road, London NW1 2BU, UK

<u>St George’s University Hospitals NHS Foundation Trust</u>

Simon B Drysdale^1^ (principal investigator)

Recruitment team; Nicole Branch^1^, Lisa Hamzah^1^, Heather Jarman^1^ (alphabetical order)

1. St George’s Hospital, Blackshaw Road, London SW17 0QT, UK

<u>Lewisham and Greenwich NHS Trust</u>

Maggie Nyirenda^1, 2^,(principal investigator)

Recruitment team Lisa Capozzi^1^, Emma Gardiner^1^ (alphabetical order)

1. University Hospital Lewisham, London SE13 6LH, UK
2. Queen Elizabeth Hospital Greenwich, London SE18 4QH, UK

<u>Liverpool University Hospitals NHS Foundation Trust</u>

Robert Moots^1^ (principal investigator); Magda Nasher^2^ (principal investigator)

Recruitment team; Anita Hanson^2^; Michelle Linforth^1^

1. Aintree University Hospital, Lower Lane, Liverpool L9 7AL, UK
2. Royal Liverpool Hospital, Prescot St, Liverpool L7 8XP, UK

<u>Leeds Teaching Hospitals NHS Trust</u>

Sean O’Riordan^1^ (principal investigator)

Recruitment team; Donna Ellis^1^

1. Leeds Children’s Hospital, Leeds LS1 3EX, UK

<u>King’s College Hospital NHS Foundation Trust</u>

Akash Deep^1^ (principal investigator)

Recruitment team; Ivan Caro^1^

1. Kings College Hospital, Denmark Hill, London SE5 9RS, UK

<u>Sheffield Children’s NHS Foundation Trust</u>

Fiona Shackley ^1^ (principal investigator);

Recruitment team; Arianna Bellini,^1^ Stuart Gormley^1^ (alphabetical order)

1. Sheffield Children’s Hospital, Broomhall, Sheffield S10 2TH, UK

<u>University Hospitals of Leicester NHS Foundation Trust</u>

Samira Neshat^1^ (principal investigator)

1. Leicester General Hospital, Leicester LE1 5WW, UK

<u>Birmingham Women’s and Children’s Hospital NHS Foundation Trust</u>

Barnaby J Scholefield^1^ (principal investigator)

Recruitment team; Ceri Robbins^1^, Helen Winmill^1^ (alphabetical order)

1. Birmingham Children’s Hospital, Steelhouse Lane, Birmingham B4 6NH, UK

**University of Oxford (UK)**

**Children’s Hospital, John Radcliffe Hospital, Oxford**

<u>Principal Investigator</u>

Stéphane C. Paulus^1,2,3^

<u>Co-Principal Investigator</u>

Andrew J. Pollard^1,2,3,4^

<u>Co-investigators</u>

Mark Anthony^1^ (neonates)

<u>Recruitment team</u>

Sarah Hopton^1^, Danielle Miller^1^, Zoe Oliver^1^, Sally Beer^1^, Bryony Ward^1^

1. John Radcliffe Hospital, Oxford University Hospitals NHS Foundation Trust, Oxford, UK
2. Department of Paediatrics, University of Oxford, UK
3. Oxford Vaccine Group, University of Oxford, UK
4. NIHR Oxford Biomedical Research Centre, Oxford, UK

**University of Oxford, Nepal Site**

<u>Principal Investigator</u>

Shrijana Shrestha^1^

<u>Co-Principal Investigator</u>

Andrew J Pollard^2,3^

**Nepal Research Team**

Meeru Gurung^1^, Puja Amatya^1^, Bhishma Pokhrel^1^, Sanjeev Man Bijukchhe^1^

**Oxford Research Team**

Tim Lubinda^2^, Sarah Kelly^2^, Peter O’Reilly^2^,

1. Paediatric Research Unit, Patan Academy of Health Sciences, Kathmandu, Nepal
2. Oxford Vaccine Group, Department of Paediatrics, University of Oxford, Oxford, United Kingdom.
3. NIHR Oxford Biomedical Research Centre, Oxford, United Kingdom.

**<u>SERGAS (Spain)</u>**

<u>Principal Investigators</u>

Federico Martinón-Torres^1^

Antonio Salas^1,2^

<u>GENVIP RESEARCH GROUP</u> (in alphabetical order):

Fernando Álvez González^1^, Xabier Bello^1,2^, Mirian Ben García^1^, Sandra Carnota^1^, Miriam Cebey-López^1^, María José Curras-Tuala^1,2^, Carlos Durán Suárez^1^, Luisa García Vicente^1^, Alberto Gómez-Carballa^1,2^, Jose Gómez Rial^1^, Pilar Leboráns Iglesias^1^, Federico Martinón-Torres^1^, Nazareth Martinón-Torres^1^, José María Martinón Sánchez^1^, Belén Mosquera Pérez^1^, Jacobo Pardo-Seco^1,2^, Lidia Piñeiro Rodríguez^1^, Sara Pischedda^1,2^, Sara Rey Vázquez^1^, Irene Rivero Calle^1^, Carmen Rodríguez-Tenreiro^1^, Lorenzo Redondo-Collazo^1^, Miguel Sadiki Ora^1^, Antonio Salas^1,2^, Sonia Serén Fernández^1^, Cristina Serén Trasorras^1^, Marisol Vilas Iglesias^1^.

1. Translational Pediatrics and Infectious Diseases, Pediatrics Department, Hospital Clínico Universitario de Santiago, Santiago de Compostela, Spain, and GENVIP Research Group (www.genvip.org), Instituto de Investigación Sanitaria de Santiago, Universidad de Santiago de Compostela, Galicia, Spain.
2. Unidade de Xenética, Departamento de Anatomía Patolóxica e Ciencias Forenses, Instituto de Ciencias Forenses, Facultade de Medicina, Universidade de Santiago de Compostela, and GenPop Research Group, Instituto de Investigaciones Sanitarias (IDIS), Hospital Clínico Universitario de Santiago, Galicia, Spain
3. Fundación Pública Galega de Medicina Xenómica, Servizo Galego de Saúde (SERGAS), Instituto de Investigaciones Sanitarias (IDIS), and Grupo de Medicina Xenómica, Centro de Investigación Biomédica en Red de Enfermedades Raras (CIBERER), Universidade de Santiago de Compostela (USC), Santiago de Compostela, Spain

**Liverpool (UK)**

<u>Principal Investigators</u>

Enitan D Carrol^1,2,3^

<u>Research Group (in alphabetical order):</u>

Elizabeth Cocklin^1^, Aakash Khanijau^1^, Rebecca Lenihan^1^, Nadia Lewis-Burke^1^, Karen Newall^4^, Sam Romaine^1^

1. Department of Clinical Infection, Microbiology and Immunology, University of Liverpool Institute of Infection, Veterinary and Ecological Sciences, Liverpool, England
2. Alder Hey Children’s Hospital, Department of Infectious Diseases, Eaton Road, Liverpool, L12 2AP
3. Liverpool Health Partners, 1st Floor, Liverpool Science Park, 131 Mount Pleasant, Liverpool, L3 5TF
4. Alder Hey Children’s Hospital, Clinical Research Business Unit, Eaton Road, Liverpool, L12 2AP

## NATIONAL AND KAPODISTRIAN UNIVERSITY OF ATHENS (Greece)

Principal Investigator: Maria Tsolia^1^

Co-Investigator: Irini Eleftheriou^1^

PID Unit: Nikos Spyridis^1^, Maria Tambouratzi^1^

Pediatric Rheumatology Unit: Despoina Maritsi^1^

Lab: Antonios Marmarinos^1^, Marietta Xagorari^1^

<u>Recruitment teams:</u>

Adult COVID19-Infectious Diseases: Lourida Panagiota, Pefanis Aggelos^2^

Adult COVID19: Akinosoglou Karolina, Gogos Charalambos, Maragos Markos^3^

Adult Inflammatory Diseases-Oncology: Voulgarelis Michalis, Stergiou Ioanna^4^

1. Department of Pediatrics, National and Kapodistrian University of Athens (NKUA), Children’s Hospital “P, and A. Kyriakou”, Athens, Greece
2. Department of Infectious Diseases, General Hospital “Sotiria”
3. Pathology Department, University of Patras, General Hospital “Panagia i Voithia”
4. Pathophysiology Department, Medical Faculty, National and Kapodistrian University of Athens (NKUA), General Hospital “Laiko”

**Newcastle upon Tyne Hospitals NHS Foundation Trust and Newcastle University (UK) combined**

<u>Principal Investigator:</u>

Marieke Emonts ^1,2,3^ (all activities)

<u>Co-investigators</u>

Emma Lim^2,3,5^ (all activities)

John Isaacs^1^ (adult inflammatory)

<u>Recruitment team (alphabetical), datamanagers, and GNCH Research unit:</u>

Kathryn Bell^4^, Stephen Crulley^4^, Daniel Fabian^4^, Evelyn Thomson^4^, Diane Wallia^4^, Caroline Miller^4^, Ashley Bell^4^

<u>PhD Students/medical staff DIAMONDS</u>

Fabian J.S. van der Velden^1,2^ (all activities), Geoff Shenton^6^ (oncology), Ashley Price^7,8^ (Adult COVID)

<u>Students</u>

Owen Treloar ^1,2^ (quality control, data management and analysis)

Daisy Thomas^1,2^ (recruitment)

<u>Author Affiliations:</u>

1. Translational and Clinical Research Institute, Newcastle University, Newcastle upon Tyne UK
2. Great North Children’s Hospital, Paediatric Immunology, Infectious Diseases & Allergy, Newcastle upon Tyne Hospitals NHS Foundation Trust, Newcastle upon Tyne, United Kingdom.
3. NIHR Newcastle Biomedical Research Centre based at Newcastle upon Tyne Hospitals NHS Trust and Newcastle University, Westgate Rd, Newcastle upon Tyne NE4 5PL, United Kingdom
4. Great North Children’s Hospital, Research Unit, Newcastle upon Tyne Hospitals NHS Foundation Trust, Newcastle upon Tyne, United Kingdom.
5. Population Health Sciences Institute, Newcastle University, Newcastle upon Tyne, UK
6. Great North Children’s Hospital, Paediatric Oncology, Newcastle upon Tyne Hospitals NHS Foundation Trust, Newcastle upon Tyne, United Kingdom.
7. Department of Infection & Tropical Medicine, Newcastle upon Tyne Hospitals NHS Foundation Trust, Newcastle upon Tyne, United Kingdom
8. NIHR Newcastle In Vitro Diagnostics Co-operative (Newcastle MIC), Newcastle upon Tyne, United Kingdom.

**<u>Servicio Madrileño de Salud (SERMAS) - Fundación Biomédica del Hospital Universitario 12 de Octubre (FIB-H12O) (Spain)</u>**

<u>Principal Investigators</u>

Pablo Rojo^1,3^

Cristina Epalza ^1,2^

<u>SERMAS/FIB-H120 team:</u>

Serena Villaverde ^1^,, Sonia Márquez^2^, Manuel Gijón ^2^, Fátima Machín^2^, Laura Cabello^2^, Irene Hernández^2^, Lourdes Gutiérrez^2^, Ángela Manzanares ^1^

<u>Author Affiliations:</u>

1. Servicio Madrileño de Salud (SERMAS),Pediatric Infectious Diseases Unit, Department of Pediatrics, Hospital Universitario 12 de Octubre, Madrid, Spain
2. Fundación Biomédica del Hospital Universitario 12 de Octubre (FIB-H12O), Unidad Pediátrica de Investigación y Ensayos Clínicos (UPIC), Hospital Universitario 12 de Octubre, Instituto de Investigación Sanitaria Hospital 12 de Octubre (i+12), Madrid, Spain.
3. Universidad Complutense de Madrid, Faculty of Medicine, Department of Pediatrics, Madrid, Spain.

**Amsterdam University Medical Center (Amsterdam UMC), University of Amsterdam**

<u>Principal Investigator:</u>

T.W. (Taco) Kuijpers MD PhD ^1,2^ (all activities)

<u>Co-investigators</u>

M. (Martijn) van de Kuip MD PhD ^1^ (infectious disease)

A.M. (Marceline) van Furth MD PhD ^1^ (infectious disease)

J.M. (Merlijn) van den Berg MD PhD ^1^ (inflammatory disease)

<u>Hospital Team (all activities):</u>

Giske Biesbroek MD PhD ^1^, Floris Verkuil MD (PhD student) ^1^, Carlijn (C.W.) van der Zee MD (start 1/8/2022, PhD student) ^1^

Recruitment:

Dasja Pajkrt MD PhD ^1^, Michael Boele van Hensbroek MD PhD ^1^, Dieneke Schonenberg MD ^1^, Mariken Gruppen MD ^1^, Sietse Nagelkerke MD PhD ^1,2^, medical students

Laboratory Team:

Machiel H Jansen ^1^, Ines Goetschalckx (PhD student) ^2^

<u>Author Affiliations:</u>

1. Amsterdam UMC, Emma Children’s Hospital, Dept of Pediatric Immunology, Rheumatology and Infectious Disease, University of Amsterdam, The Netherlands
2. Sanquin, Dept of Molecular Hematology, University Medical Center, Amsterdam, The Netherlands

**Bambino Gesù Children’s Hospital (Rome-Italy)**

<u>Principal Investigator</u>

Lorenza Romani 1

Maia De Luca 1

<u>Recruitment Team</u>

Sara Chiurchiù 1

Martina Di Giuseppe 1

<u>Affiliation</u>

1. Infectious Disease Unit, Academic Department of Pediatrics, Bambino Gesù Children’s Hospital, IRCCS, Rome 00165, Italy

**ERASMUS MC-Sophia Children’s Hospital**

Principal Investigator

Clementien L. Vermont²

Research group

Henriëtte A. Moll¹, Dorine M. Borensztajn¹, Nienke N. Hagedoorn, Chantal Tan ¹, Joany Zachariasse ¹, Medical students ¹

<u>Additional investigator</u>

W Dik ^3^

1. Erasmus MC-Sophia Children’s Hospital, Department of General Paediatrics, Rotterdam, the Netherlands
2. Erasmus MC-Sophia Children’s Hospital, Department of Paediatric Infectious Diseases & Immunology, Rotterdam, the Netherlands
3. Erasmus MC, Department of immunology, Rotterdam, the Netherlands

**TAIWAN**

Ching-Fen (Kitty), Shen^1^

1. Division of Infectious Disease, Department of Pediatrics, National Cheng Kung University, Tainan, Taiwan

**Riga Stradins University (Riga, Latvia)**

<u>Principal Investigator:</u>

Dace Zavadska ^1,2^ (all activities)

<u>Co-investigators</u>

Sniedze Laivacuma ^1,3^ (adult cohorts)

<u>Recruitment team:</u>

Aleksandra Rudzate ^1,2^, Diana Stoldere ^1,2^, Arta Barzdina ^1,2^, Elza Barzdina ^1,2^, Sniedze Laivacuma^1,3^, Monta Madelane ^1,3^

<u>Laboratory</u>

Dagne Gravele ^2^, Dace Svile^2^

<u>Author Affiliations:</u>

1. Riga Stradins University, Riga, Latvia
2. Children clinical university hospital, Riga, Latvia
3. Riga East clinical university hospital, Riga, Latvia

**Assistance Publique - Hôpitaux de Paris**

<u>Principal Investigator:</u>

Romain Basmaci ^1,2^

<u>Co-investigator:</u>

Noémie Lachaume ^1^

Recruitment team:

Pauline Bories ^1^, Raja Ben Tkhayat ^1^, Laura Chériaux ^1^, Juraté Davoust ^1^, Kim-Thanh Ong ^1^, Marie Cotillon ^1^, Thibault de Groc ^1^, Sébastien Le ^1^, Nathalie Vergnault ^1^, Hélène Sée ^1^, Laure Cohen ^1^, Alice de Tugny ^1^, Nevena Danekova ^1^

<u>Author Affiliations:</u>

1. Service de Pédiatrie-Urgences, AP-HP, Hôpital Louis-Mourier, F-92700 Colombes, France
2. Université Paris Cité, Inserm, IAME, F-75018 Paris, France

**BioMérieux**

<u>Principal Investigator:</u>

Marine Mommert-Tripon^1^

<u>Co-investigator:</u>

Karen Brengel-Pesce^1^

<u>Author Affiliations:</u>

1. bioMérieux - Open Innovation & Partnerships Department, Lyon, France

**University Medical Centre Ljubljana, Slovenia**

Principal Investigator: Marko Pokorn ^1,2,3^

Co-Investigator: Mojca Kolnik^2^

Research Group (in alphabetical order):

Tadej Avčin^2,3^, Tanja Avramoska^2^, Natalija Bahovec^1^, Petra Bogovič^1^, Lidija Kitanovski^2,3^, Mirijam Nahtigal^1^, Lea Papst^1^, Tina Plankar Srovin^1^, Franc Strle^1,2^, Anja Srpčič^2^, Katarina Vincek^1^.

Affiliations:

1. Department of Infectious diseases, University Medical Centre Ljubljana, Slovenia
2. University Children’s Hospital, University Medical Centre Ljubljana, Slovenia
3. Faculty of Medicine, University of Ljubljana, Slovenia
4. Centre for Clinical research, University Medical Centre Ljubljana

**University Medical Center Utrecht, Utrecht, The Netherlands**

<u>Principal Investigator</u>

Michiel van der Flier^1,5^ ^(^Pediatric Infectious Diseases and Immunology)

<u>Co-investigators</u>

Wim J.E. Tissing^5^ (Pediatric Oncology)

Roelie M. Wösten-van Asperen^2^ (Pediatric Intensive Care Unit)

Sebastiaan J Vastert^3^ (Pediatric Rheumatology)

Daniel C Vijlbrief^4^ (Pediatric Neonatal Intensive Care)

Louis J. Bont^1,5^ ^(^Pediatric Infectious Diseases and Immunology)

Tom F.W. Wolfs ^1,5^ ^(^Pediatric Infectious Diseases and Immunology)

<u>PhD student</u>

Coco R. Beudeker^1,5^ ^(^Pediatric Infectious Diseases and Immunology)

<u>Affiliations:</u>

1. Pediatric Infectious Diseases and Immunology,
2. Pediatric Intensive Care Unit
3. Pediatric Rheumatology
4. Pediatric Neonatal Intensive Care, Wilhelmina Children’s Hospital, University Medical Center Utrecht, Utrecht, The Netherlands
5. Princess Maxima Center for Pediatric Oncology, Utrecht, The Netherlands

**University of Bern, Inselspital, Bern University Hospital, University of Bern (Switzerland)**

<u>Principal Investigators (alphabetical)</u>

Philipp Agyeman^1^

Luregn Schlapbach^2,3^

<u>Co-Investigator</u>

Christoph Aebi^1^

<u>Recruitment team</u>

Mariama Usman^1^, Stefanie Schlüchter^1^, Verena Wyss^1^, Nina Schöbi^1^, Elisa Zimmermann^2^ PhD, Marion Meier^2^, Kathrin Weber^2^

1. Department of Pediatrics, Inselspital, Bern University Hospital, University of Bern, Switzerland
2. Department of Intensive Care and Neonatology, and Children’s Research Center, University Children’s Hospital Zurich, Zurich, Switzerland
3. Child Health Research Centre, The University of Queensland, Brisbane, Australia

**Swiss Pediatric Sepsis Study group**

Philipp Agyeman, MD ^1^, Luregn J Schlapbach, MD, FCICM ^2,3^, Eric Giannoni, MD ^4,5^, Martin Stocker, MD ^6^, Klara M Posfay-Barbe, MD ^7^, Ulrich Heininger, MD ^8^, Sara Bernhard-Stirnemann, MD ^9^, Anita Niederer-Loher, MD ^10^, Christian Kahlert, MD ^10^, Giancarlo Natalucci, MD ^11^, Christa Relly, MD ^12^, Thomas Riedel, MD ^13^, Christoph Aebi, MD ^1^, Christoph Berger, MD ^12^

**Affiliations:**

1. Department of Pediatrics, Inselspital, Bern University Hospital, University of Bern, Switzerland
2. Department of Intensive Care and Neonatology, and Children’s Research Center, University Children’s Hospital Zurich, Zurich, Switzerland
3. Child Health Research Centre, The University of Queensland, Brisbane, Australia
4. Clinic of Neonatology, Department Mother-Woman-Child, Lausanne University Hospital and University of Lausanne, Switzerland
5. Infectious Diseases Service, Department of Medicine, Lausanne University Hospital and University of Lausanne, Switzerland
6. Department of Pediatrics, Children’s Hospital Lucerne, Lucerne, Switzerland
7. Pediatric Infectious Diseases Unit, Children’s Hospital of Geneva, University Hospitals of Geneva, Geneva, Switzerland
8. Infectious Diseases and Vaccinology, University of Basel Children’s Hospital, Basel, Switzerland
9. Children’s Hospital Aarau, Aarau, Switzerland
10. Division of Infectious Diseases and Hospital Epidemiology, Children’s Hospital of Eastern Switzerland St. Gallen, St. Gallen, Switzerland
11. Department of Neonatology, University Hospital Zurich, Zurich, Switzerland
12. Division of Infectious Diseases and Hospital Epidemiology, and Children’s Research Center, University Children’s Hospital Zurich, Switzerland
13. Children’s Hospital Chur, Chur, Switzerland

**Micropathology Ltd (UK)**

Principal Investigator; Prof Colin Fink^1^

Co Investigators: Marie Voice^1^, Leo Calvo-Bado^1^, Michael Steele^1^, Jennifer Holden^1^ Research group: Benjamin Evans^1^, Jake Stevens^1^, Peter Matthews^1^, Kyle Billing^1^

1. Micropathology Ltd, The Venture Center, University of Warwick Science Park, Sir William Lyons Road, Coventry, CV4 7EZ

**Medical University of Graz, Austria (MUG)**

<u>Principal Investigator:</u>

Werner Zenz^1^ (all activities)

<u>Co-investigators (in alphabetical order):</u>

Alexander Binder^1^ (grant application)

Benno Kohlmaier^1^ (study design, recruitment)

Daniela S. Kohlfürst^1^ (study design)

Nina A. Schweintzger^1^ (all activities)

Christoph Zurl^1^ (study design, recruitment)

<u>Recruitment team, data managers, laboratory work (in alphabetical order):</u>

Susanne Hösele^1^, Manuel Leitner^1^, Lena Pölz^1^, Alexandra Rusu^1^, Glorija Rajic^1^, Bianca Stoiser^1^, Martina Strempfl^1^, Manfred G. Sagmeister^1^

<u>Clinical recruitment partners (in alphabetical order):</u>

Sebastian Bauchinger^1^, Martin Benesch^3^, Astrid Ceolotto^1^, Ernst Eber^2^, Siegfried Gallistl^1^, Harald Haidl^1^, Almuthe Hauer^1^, Christa Hude^1^, Andreas Kapper^7^, Markus Keldorfer^5^, Sabine Löffler^5^, Tobias Niedrist^6^, Heidemarie Pilch^5^, Andreas Pfleger^2^, Klaus Pfurtscheller^4^, Siegfried Rödl^4^, Andrea Skrabl-Baumgartner^1^, Volker Strenger^3^, Elmar Wallner^7^

<u>Author Affiliations:</u>

1. Department of Pediatrics and Adolescent Medicine, Division of General Pediatrics, Medical University of Graz, Graz, Austria
2. Department of Pediatric Pulmonology, Medical University of Graz, Graz, Austria
3. Department of Pediatric Hematooncology, Medical University of Graz, Graz, Austria
4. Paediatric Intensive Care Unit, Medical University of Graz, Graz, Austria
5. University Clinic of Pediatrics and Adolescent Medicine Graz, Medical University Graz, Graz, Austria
6. Clinical Institute of Medical and Chemical Laboratory Diagnostics, Medical University Graz, Graz, Austria
7. Department of Internal Medicine, State Hospital Graz II, Location West, Graz, Austria

**SkylineDX**

Principle investigator: Dennie Tempel ^1^

Co-investigators: Danielle van Keulen^1^, Annelieke M Strijbosch ^1^,

Author affiliations:

1. SkylineDx, Rotterdam, The Netherlands

**BBMRI-ERIC**

Maike K. Tauchert^1^ Author affiliation:

1. Biobanking and BioMolecular Resources Research Infrastructure – European Research Infrastructure Consortium (BBMRI-ERIC), Neue Stiftingtalstrasse 2/B/6, 8010, Graz, Austria

**LMU Munich Partner (Germany)**

<u>Principal Investigator:</u>

Ulrich von Both^1,2^ MD, FRCPCH (all activities)

<u>Research group:</u>

Laura Kolberg¹ MSc (all activities)

Patricia Schmied¹ (Study physician), Irene Alba-Alejandre^3^ MD (Study physician)

<u>Clinical recruitment partners (in alphabetical order):</u>

Katharina Danhauser, MD^6^, Nikolaus Haas, MD^11^, Florian Hoffmann, MD^10^, Matthias Griese, MD^7^, Tobias Feuchtinger, MD^5^, Sabrina Juranek, MD^4^, Matthias Kappler, MD^7^, Eberhard Lurz, MD^8^, Esther Maier, MD^4^, Karl Reiter, MD^10^, Carola Schoen, MD^10^, Sebastian Schroepf, MD^9^

<u>Author Affiliations:</u>

1. Division of Pediatric Infectious Diseases, Department of Pediatrics, Dr. von Hauner Children’s Hospital, University Hospital, LMU Munich, Munich, Germany
2. German Center for Infection Research (DZIF), Partner Site Munich, Munich, Germany
3. Department of Gynecology and Obstetrics, University Hospital, LMU Munich, Munich, Germany
4. Division of General Pediatrics, Department of Pediatrics, Dr. von Hauner Children’s Hospital, University Hospital, LMU Munich, Munich, Germany
5. Division of Pediatric Haematology & Oncology, Department of Pediatrics, Dr. von Hauner Children’s Hospital, University Hospital, LMU Munich, Munich, Germany
6. Division of Pediatric Rheumatology, Department of Pediatrics, Dr. von Hauner Children’s Hospital, University Hospital, LMU Munich, Munich, Germany
7. Division of Pediatric Pulmonology, Department of Pediatrics, Dr. von Hauner Children’s Hospital, University Hospital, LMU Munich, Munich, Germany
8. Division of Pediatric Gastroenterology, Department of Pediatrics, Dr. von Hauner Children’s Hospital, University Hospital, LMU Munich, Munich, Germany
9. Neonatal Intensive Care Unit, Department of Pediatrics, Dr. von Hauner Children’s Hospital, University Hospital, LMU Munich, Munich, Germany
10. Paediatric Intensive Care Unit, Department of Pediatrics, Dr. von Hauner Children’s Hospital, University Hospital, LMU Munich, Munich, Germany
11. Department of Pediatric Cardiology and Pediatric Intensive Care, University Hospital, LMU Munich, Germany

**London School of Hygiene and Tropical Medicine (LSHTM)**

<u>Principal Investigator: Shunmay Yeung</u> ^1,2,3^

Research group:

Manuel Dewez^1^ David Bath^3^, Elizabeth Fitchett^1^, Fiona Cresswell^1^

1. Clinical Research Department, Faculty of Infectious and Tropical Disease, London School of Hygiene and Tropical Medicine, London
2. 1. ^2^. Department of Paediatrics, St. Mary’s Imperial College Hospital, London
3. Department of Global Health and Development, Faculty of Public Health and Policy

**PERFORM Consortium**

https://www.perform2020.org/

**<u>PARTNER: IMPERIAL COLLEGE (UK)</u>**

<u>Chief investigator/PERFORM coordinator:</u>

Michael Levin

<u>Principal and co-investigators; work package leads (alphabetical order)</u>

Aubrey Cunnington (grant application)

Tisham De (work package lead)

Jethro Herberg (Principle Investigator, Deputy Coordinator, grant application)

Myrsini Kaforou (grant application, work package lead)

Victoria Wright (grant application, Scientific Coordinator)

<u>Research Group (alphabetical order)</u>

Lucas Baumard; Evangelos Bellos; Giselle D’Souza; Rachel Galassini; Dominic Habgood-Coote; Shea Hamilton; Clive Hoggart; Sara Hourmat; Heather Jackson; Ian Maconochie; Stephanie Menikou; Naomi Lin; Samuel Nichols; Ruud Nijman; Ivonne Pena Paz; Oliver Powell, Priyen Shah; Ching-Fen Shen; Clare Wilson

<u>Clinical recruitment at Imperial College Healthcare NHS Trust (alphabetical order)</u>

Amina Abdulla; Ladan Ali; Sarah Darnell; Rikke Jorgensen; Sobia Mustafa; Salina Persand

<u>Imperial College Faculty of Engineering</u>

Molly Stevens (co-investigator), Eunjung Kim (research group); Benjamin Pierce (research group)

<u>Clinical recruitment at Brighton and Sussex University Hospitals</u>

Katy Fidler (Principle Investigator)

Julia Dudley (Clinical Research Registrar)

Research nurses: Vivien Richmond, Emma Tavliavini

<u>Clinical recruitment at National Cheng Kung University Hospital</u>

Ching-Fen Shen (Principal Investigator); Ching-Chuan Liu (Co-investigator); Shih-Min Wang (Co-investigator), funded by the Center of Clinical Medicine Research, National Cheng Kung University

**<u>PARTNER: SERGAS (Spain)</u>**

<u>Principal Investigators</u>

Federico Martinón-Torres^1^

Antonio Salas^1,2^

<u>Research Group (alphabetical order)</u>

Fernando Álvez González^1^, Cristina Balo Farto^1^, Ruth Barral-Arca^1,2^, María Barreiro Castro^1^, Xabier Bello^1,2^, Mirian Ben García^1^, Sandra Carnota^1^, Miriam Cebey-López^1^, María José Curras-Tuala^1,2^, Carlos Durán Suárez^1^, Luisa García Vicente^1^, Alberto Gómez-Carballa^1,2^, Jose Gómez Rial^1^, Pilar Leboráns Iglesias^1^, Federico Martinón-Torres^1^, Nazareth Martinón-Torres^1^, José María Martinón Sánchez^1^, Belén Mosquera Pérez^1^, Jacobo Pardo-Seco^1,2^, Lidia Piñeiro Rodríguez^1^, Sara Pischedda^1,2^, Sara Rey Vázquez^1^, Irene Rivero Calle^1^, Carmen Rodríguez-Tenreiro^1^, Lorenzo Redondo-Collazo^1^, Miguel Sadiki Ora^1^, Antonio Salas^1,2^, Sonia Serén Fernández^1^, Cristina Serén Trasorras^1^, Marisol Vilas Iglesias^1^.

**<u>PARTNER: RSU (Latvia)</u>**

<u>Principal Investigator</u>

Dace Zavadska^1,2^

<u>Other RSU group authors (in alphabetical order):</u>

Anda Balode^1,2^, Arta Bārzdiņa^1,2^, Dārta Deksne^1,2^, Dace Gardovska^1,2^, Dagne Grāvele^2^, Ilze Grope^1,2^, Anija Meiere^1,2^, Ieva Nokalna^1,2^, Jana Pavāre^1,2^, Zanda Pučuka^1,2^, Katrīna Selecka^1,2^, Aleksandra Sidorova^1,2^, Dace Svile^2^, Urzula Nora Urbāne^1,2^.

1. Riga Stradins university, Riga, Latvia.
2. Children clinical university hospital, Riga, Latvia.

**<u>PARTNER: Medical Research Council Unit The Gambia (MRCG) at LSHTM</u>**

<u>Principal Investigator</u>

Effua Usuf^1^

<u>Additional Investigators</u>

Kalifa Bojang^1^

Syed M. A. Zaman^1^

Fatou Secka^1^

Suzanne Anderson^1^

Anna RocaIsatou Sarr^1^

Momodou Saidykhan^1^

Saffiatou Darboe^1^

Samba Ceesay^1^

Umberto D’alessandro^1^

1. Medical Research Council Unit The Gambia at LSHTM, P O Box 273, Fajara, The Gambia

**<u>ERASMUS MC-Sophia Children’s Hospital (Netherlands</u>**

<u>Principal Investigator</u>

Henriëtte A. Moll¹

<u>Research Group (alphabetical order)</u>

Dorine M. Borensztajn¹, Nienke N. Hagedoorn, Chantal Tan ¹, ¹, Clementien L. Vermont², Joany Zachariasse ¹

<u>Additional investigator</u>

W Dik ^3^

1. Erasmus MC-Sophia Children’s Hospital, Department of General Paediatrics, Rotterdam, the Netherlands
2. Erasmus MC-Sophia Children’s Hospital, Department of Paediatric Infectious Diseases & Immunology, Rotterdam, the Netherlands
3. Erasmus MC, Department of immunology, Rotterdam, the Netherlands

**<u>Swiss Pediatric Sepsis Study (Switzerland)</u>**

<u>Principal Investigators:</u>

Philipp Agyeman, MD ^1^ (ORCID 0000-0002-8339-5444), Luregn J Schlapbach, MD, FCICM ^2,3^ (ORCID 0000-0003-2281-2598)

<u>Clinical recruitment at University Children’s Hospital Bern for PERFORM:</u>

Christoph Aebi ^1^, Verena Wyss ^1^, Mariama Usman ^1^

<u>Principal and co-investigators for the Swiss Pediatric Sepsis Study:</u>

Philipp Agyeman, MD ^1^, Luregn J Schlapbach, MD, FCICM ^2,3^, Eric Giannoni, MD ^4,5^, Martin Stocker, MD ^6^, Klara M Posfay-Barbe, MD ^7^, Ulrich Heininger, MD ^8^, Sara Bernhard-Stirnemann, MD ^9^, Anita Niederer-Loher, MD ^10^, Christian Kahlert, MD ^10^, Giancarlo Natalucci, MD ^11^, Christa Relly, MD ^12^, Thomas Riedel, MD ^13^, Christoph Aebi, MD ^1^, Christoph Berger, MD ^12^ for the Swiss Pediatric Sepsis Study

1. Department of Pediatrics, Inselspital, Bern University Hospital, University of Bern, Switzerland
2. Neonatal and Pediatric Intensive Care Unit, Children’s Research Center, University Children’s Hospital Zurich, University of Zurich, Zurich, Switzerland
3. Child Health Research Centre, University of Queensland, and Queensland Children’s Hospital, Brisbane, Australia
4. Clinic of Neonatology, Department Mother-Woman-Child, Lausanne University Hospital and University of Lausanne, Switzerland
5. Infectious Diseases Service, Department of Medicine, Lausanne University Hospital and University of Lausanne, Switzerland
6. Department of Pediatrics, Children’s Hospital Lucerne, Lucerne, Switzerland
7. Pediatric Infectious Diseases Unit, Children’s Hospital of Geneva, University Hospitals of Geneva, Geneva, Switzerland
8. Infectious Diseases and Vaccinology, University of Basel Children’s Hospital, Basel, Switzerland
9. Children’s Hospital Aarau, Aarau, Switzerland
10. Division of Infectious Diseases and Hospital Epidemiology, Children’s Hospital of Eastern Switzerland St. Gallen, St. Gallen, Switzerland
11. Department of Neonatology, University Hospital Zurich, Zurich, Switzerland
12. Division of Infectious Diseases and Hospital Epidemiology, and Children’s Research Center, University Children’s Hospital Zurich, Switzerland
13. Children’s Hospital Chur, Chur, Switzerland

**<u>Liverpool (UK)</u>**

<u>Principal Investigators</u>

Enitan D Carrol^1,2,3^

Stéphane Paulus ^1^

<u>Research Group (alphabetical order)</u>

Elizabeth Cocklin^1^, Rebecca Jennings^4^, Joanne Johnston^4^, Simon Leigh^1^, Karen Newall^4^, Sam Romaine^1^

1. Department of Clinical Infection, Microbiology and Immunology, University of Liverpool Institute of Infection and Global Health, Liverpool, England
2. Alder Hey Children’s Hospital, Department of Infectious Diseases, Eaton Road, Liverpool, L12 2AP
3. Liverpool Health Partners, 1^st^ Floor, Liverpool Science Park, 131 Mount Pleasant, Liverpool, L3 5TF
4. Alder Hey Children’s Hospital, Clinical Research Business Unit, Eaton Road, Liverpool, L12 2AP

**<u>NKUA (Greece)</u>**

<u>Principal investigator</u>

Professor Maria Tsolia^1^ (all activities)

<u>Investigator/Research fellow</u>

Irini Eleftheriou^1^ (all activities)

<u>Additional investigators</u>

Recruitment: Maria Tambouratzi^1^

Lab: Antonis Marmarinos^1^ (Quality Manager)

Lab: Marietta Xagorari^1^

Kelly Syggelou^1^

1. Department of Pediatrics, National and Kapodistrian University of Athens, “P. and A. Kyriakou” Children’s Hospital Thivon and Levadias Goudi, Athens

**<u>Micropathology Ltd (UK)</u>**

<u>Principal Investigator</u>

Professor Colin Fink^1^, Clinical Microbiologist

<u>Additional investigators</u>

Dr Marie Voice^1^, Post doc scientist

Dr. Leo Calvo-Bado^1^, Post doc scientist

1. Micropathology Ltd, The Venture Center, University of Warwick Science Park, Sir William Lyons Road, Coventry, CV4 7EZ

**<u>Medical University of Graz (MUG, Austria)</u>**

<u>Principal Investigator</u>

Werner Zenz^1^ (all activities)

<u>Co-investigators (alphabetical order)</u>

Benno Kohlmaier^1^ (all activities)

Nina A. Schweintzger^1^ (all activities)

Manfred G. Sagmeister^1^ (study design, consortium wide sample management)

<u>Research team</u>

Daniela S. Kohlfürst^1^ (study design)

Christoph Zurl^1^ (BIVA PIC)

Alexander Binder^1^ (grant application)

<u>Recruitment team, data managers, (alphabetical order)</u>

Susanne Hösele^1^, Manuel Leitner^1^, Lena Pölz^1^, Glorija Rajic^1^,

<u>Clinical recruitment partners (alphabetical order)</u>

Sebastian Bauchinger^1^, Hinrich Baumgart^4^, Martin Benesch^3^, Astrid Ceolotto^1^, Ernst Eber^2^, Siegfried Gallistl^1^, Gunther Gores^5^, Harald Haidl^1^, Almuthe Hauer^1^, Christa Hude^1^, Markus Keldorfer^5^, Larissa Krenn^4^, Heidemarie Pilch^5^, Andreas Pfleger^2^, Klaus Pfurtscheller^4^, Gudrun Nordberg^5^, Tobias Niedrist^8^, Siegfried Rödl^4^, Andrea Skrabl-Baumgartner^1^, Matthias Sperl^7^, Laura Stampfer^5^, Volker Strenger^3^, Holger Till^6^, Andreas Trobisch^5^, Sabine Löffler^5^

1. Department of Pediatrics and Adolescent Medicine, Division of General Pediatrics, Medical University of Graz, Graz, Austria
2. Department of Pediatric Pulmonology, Medical University of Graz, Graz, Austria
3. Department of Pediatric Hematooncoloy, Medical University of Graz, Graz, Austria
4. Paediatric Intensive Care Unit, Medical University of Graz, Graz, Austria
5. University Clinic of Paediatrics and Adolescent Medicine Graz, Medical University Graz, Graz,Austria
6. Department of Paediatric and Adolescence Surgery, Medical University Graz, Graz, Austria
7. Department of Pediatric Orthopedics, Medical University Graz, Graz, Austria
8. Clinical Institute of Medical and Chemical Laboratory Diagnostics, Medical University Graz, Graz, Austria

**<u>London School of Hygiene and Tropical Medicine (UK)</u>**

<u>Principal Investigator:</u>

Dr Shunmay Yeung^1,2^ ^3^ PhD, MBBS, FRCPCH, MRCP, DTM&H

<u>Research Group</u>

Dr Juan Emmanuel Dewez^1^ MD, DTM&H, MSc

Prof Martin Hibberd ^1^ BSc, PhD

Mr David Bath^2^ MSc, MappFin, BA(Hons)

Dr Alec Miners^2^ BA(Hons), MSc, PhD

Dr Ruud Nijman^3^ PhD MSc MD MRCPCH

Dr Catherine Wedderburn^1^ BA, MBChB, DTM&H, MSc, MRCPCH

Ms Anne Meierford^1^ MSc, BmedSc, BMBS

Dr Baptiste Leurent^4^, PhD, MSc

1. Faculty of Infectious and Tropical Disease, London School of Hygiene and Tropical Medicine, London, UK
2. Faculty of Public Health and Policy, London School of Hygiene and Tropical Medicine, London, UK
3. Department of Paediatrics, St. Mary’s Hospital Imperial College Hospital, London, UK
4. Faculty of Epidemiology and Population Health, London School of Hygiene and Tropical Medicine, London, UK

**<u>Radboud University Medical Center (RUMC, Netherlands)</u>**

<u>Principal Investigators</u>

Ronald de Groot ^1^, Michiel van der Flier ^1,2,3^, Marien I. de Jonge^1^

<u>Co-investigators Radboud University Medical Center (alphabetical order)</u>

Koen van Aerde^1,2^, Wynand Alkema^1^, Bryan van den Broek^1^, Jolein Gloerich^1^, Alain J. van Gool^1^, Stefanie Henriet^1,2^, Martijn Huijnen^1^, Ria Philipsen^1^, Esther Willems^1^

<u>Investigators PeDBIG PERFORM DUTCH CLINICAL NETWORK (alphabetical order)</u>

G.P.J.M. Gerrits^8^, M. van Leur^8^, J. Heidema ^4^,L. de Haan^1,2^ C.J. Miedema ^5^, C. Neeleman ^1^ C.C. Obihara ^6^, G.A. Tramper-Stranders7^6^

1. Radboud University Medical Center, Nijmegen, The Netherlands
2. Amalia Children’s Hospital, Nijmegen, The Netherlands
3. Wilhelmina Children’s Hospital, University Medical Center Utrecht, Utrecht, The Netherlands
4. St. Antonius Hospital, Nieuwegein, The Netherlands
5. Catharina Hospital, Eindhoven, The Netherlands
6. ETZ Elisabeth, Tilburg, The Netherlands
7. Franciscus Gasthuis, Rotterdam, The Netherlands
8. Canisius Wilhelmina Hospital, Nijmegen, The Netherlands

**<u>Oxford (UK)</u>**

<u>Principal Investigators</u>

Andrew J. Pollard^1,2^, Rama Kandasamy^1,2^, Stéphane Paulus ^1,2^

<u>Additional Investigators</u>

Michael J. Carter^1,2^, Daniel O’Connor^1,2^, Sagida Bibi^1,2^, Dominic F. Kelly^1,2^, Meeru Gurung^3^, Stephen Thorson^3^, Imran Ansari^3^, David R. Murdoch^4^, Shrijana Shrestha^3^.

1. Oxford Vaccine Group, Department of Paediatrics, University of Oxford, Oxford, United Kingdom.
2. NIHR Oxford Biomedical Research Centre, Oxford, United Kingdom.
3. Paediatric Research Unit, Patan Academy of Health Sciences, Kathmandu, Nepal.
4. Department of Pathology, University of Otago, Christchurch, New Zealand.

**<u>Newcastle University, Newcastle upon Tyne, (UK)</u>**

<u>Principal Investigator</u>

Marieke Emonts ^1,2,3^ (all activities)

<u>Co-investigators</u>

Emma Lim^2,3,7^ (all activities) Lucille Valentine^4^

<u>Recruitment team (alphabetical), data-managers, and GNCH Research unit</u>

Karen Allen^5^, Kathryn Bell^5^, Adora Chan^5^, Stephen Crulley^5^, Kirsty Devine^5^, Daniel Fabian^5^, Sharon King^5^, Paul McAlinden^5^, Sam McDonald^5^, Anne McDonnell2,^5^, Ailsa Pickering^2,5^, Evelyn Thomson^5^, Amanda Wood^5^, Diane Wallia^5^, Phil Woodsford^5^, Sample processing: Frances Baxter^5^, Ashley Bell^5^, Mathew Rhodes^5^

<u>PICU recruitment</u>

Rachel Agbeko^8^

Christine Mackerness^8^

<u>Students MOFICHE</u>

Bryan Baas^2^, Lieke Kloosterhuis^2^, Wilma Oosthoek^2^

<u>Students/medical staff PERFORM</u>

Tasnim Arif^6^, Joshua Bennet^2^, Kalvin Collings^2^, Ilona van der Giessen^2^, Alex Martin^2^, Aqeela Rashid^6^, Emily Rowlands^2^, Gabriella de Vries^2^, Fabian van der Velden^2^

<u>Engagement work/ethics/cost effectiveness</u>

Lucille Valentine ^4^, Mike Martin^9^, Ravi Mistry^2^, Lucille Valentine^4^

1. Translational and Clinical Research Institute, Newcastle University, Newcastle upon Tyne UK
2. Great North Children’s Hospital, Paediatric Immunology, Infectious Diseases & Allergy, Newcastle upon Tyne Hospitals NHS Foundation Trust, Newcastle upon Tyne, United Kingdom.
3. NIHR Newcastle Biomedical Research Centre based at Newcastle upon Tyne Hospitals NHS Trust and Newcastle University, Westgate Rd, Newcastle upon Tyne NE4 5PL, United Kingdom
4. Newcastle University Business School, Centre for Knowledge, Innovation, Technology and Enterprise (KITE), Newcastle upon Tyne, United Kingdom
5. Great North Children’s Hospital, Research Unit, Newcastle upon Tyne Hospitals NHS Foundation Trust, Newcastle upon Tyne, United Kingdom.
6. Great North Children’s Hospital, Paediatric Oncology, Newcastle upon Tyne Hospitals NHS Foundation Trust, Newcastle upon Tyne, United Kingdom.
7. Population Health Sciences Institute, Newcastle University, Newcastle upon Tyne, UK
8. Great North Children’s Hospital, Paediatric Intensive Care Unit, Newcastle upon Tyne Hospitals NHS Foundation Trust, Newcastle upon Tyne, United Kingdom.
9. Northumbria University, Newcastle upon Tyne, United Kingdom.

**<u>LMU Munich (Germany)</u>**

<u>Principal Investigator</u>

Ulrich von Both^1,2^ MD, FRCPCH (all activities)

<u>Research group</u>

Laura Kolberg¹ MSc (all activities)

Manuela Zwerenz¹ MSc, Judith Buschbeck¹ PhD

<u>Clinical recruitment partners (alphabetical order)</u>

Christoph Bidlingmaier^3^, Vera Binder^4^, Katharina Danhauser^5^, Nikolaus Haas^10^, Matthias Griese^6^, Tobias Feuchtinger^4^, Julia Keil^9^, Matthias Kappler^6^, Eberhard Lurz^7^, Georg Muench^8^, Karl Reiter^9^, Carola Schoen^9^

1. Div. Paediatric Infectious Diseases, Hauner Children’s Hospital, University Hospital, Ludwig Maximilians University (LMU), Munich, Germany
2. German Center for Infection Research (DZIF), Partner Site Munich, Munich, Germany
3. Div. of General Paediatrics, Hauner Children’s Hospital, University Hospital, Ludwig Maximilians University (LMU), Munich, Germany
4. Div. Paediatric Haematology & Oncology, Hauner Children’s Hospital, University Hospital, Ludwig Maximilians University (LMU), Munich, Germany
5. Div. of Paediatric Rheumatology, Hauner Children’s Hospital, University Hospital, Ludwig Maximilians University (LMU), Munich, Germany
6. Div. of Paediatric Pulmonology, Hauner Children’s Hospital, University Hospital, Ludwig Maximilians University (LMU), Munich, Germany
7. Div. of Paediatric Gastroenterology, Hauner Children’s Hospital, University Hospital, Ludwig Maximilians University (LMU), Munich, Germany
8. Neonatal Intensive Care Unit, Hauner Children’s Hospital, University Hospital, Ludwig Maximilians University (LMU), Munich, Germany
9. Paediatric Intensive Care Unit Hauner Children’s Hospital, University Hospital, Ludwig Maximilians University (LMU), Munich, Germany,
10. Department Pediatric Cardiology and Pediatric Intensive Care, University Hospital, Ludwig Maximilians University (LMU), Munich, Germany

**<u>BioMérieux (France)</u>**

<u>Principal Investigator</u>

François Mallet^1,2, 3^

<u>Research Group</u>

Karen Brengel-Pesce^1,2, 3^

Alexandre Pachot^1^

Marine Mommert^1,2^

1. Open Innovation & Partnerships (OIP), bioMérieux S.A., Marcy l’Etoile, France
2. Joint research unit Hospice Civils de Lyon – bioMérieux, Centre Hospitalier Lyon Sud, 165 Chemin du Grand Revoyet, 69310 Pierre-Bénite, France
3. EA 7426 Pathophysiology of Injury-induced Immunosuppression, University of Lyon1-Hospices Civils de Lyon-bioMérieux, Hôpital Edouard Herriot, 5 Place d’Arsonval, 69437 Lyon Cedex 3, France

**<u>University Medical Centre Ljubljana (Slovenia)</u>**

<u>Principal Investigator</u>

Marko Pokorn^1,2,3^ MD, PhD

<u>Research Group</u>

Mojca Kolnik^1^ MD, Katarina Vincek^1^ MD, Tina Plankar Srovin^1^ MD, PhD, Natalija Bahovec^1^ MD, Petra Prunk^1^ MD, Veronika Osterman^1^ MD, Tanja Avramoska^1^ MD

1. Department of Infectious Diseases, University Medical Centre Ljubljana, Japljeva 2, SI-1525 Ljubljana, Slovenia
2. University Childrens’ Hospital, University Medical Centre Ljubljana, Ljubljana, Slovenia
3. Department of Infectious Diseases and Epidemiology, Faculty of Medicine, University of Ljubljana, Slovenia

**<u>Amsterdam, Academic Medical Hospital & Sanquin Research Institute (Netherlands)</u>**

<u>Principal Investigator</u>

Taco Kuijpers ^1,2^

<u>Co-investigators</u>

Ilse Jongerius ^2^

<u>Recruitment team (EUCLIDS, PERFORM)</u>

J.M. van den Berg^1^, D. Schonenberg^1^, A.M. Barendregt^1^, D. Pajkrt^1^, M. van der Kuip^1,3^, A.M. van Furth^1,3^

<u>Students PERFORM</u>

Evelien Sprenkeler ^2^, Judith Zandstra ^2^

<u>Technical support PERFORM</u>

1. G. van Mierlo ^2^, J. Geissler ^2^
2. Amsterdam University Medical Center (Amsterdam UMC), location Academic Medical Center (AMC), Dept of Pediatric Immunology, Rheumatology and Infectious Diseases, University of Amsterdam, Amsterdam, the Netherlands
3. Sanquin Research Institute, & Landsteiner Laboratory at the AMC, University of Amsterdam, Amsterdam, the Netherlands.
4. Amsterdam University Medical Center (Amsterdam UMC), location Vrije Universiteit Medical Center (VUMC), Dept of Pediatric Infectious Diseases and Immunology, Free University (VU), Amsterdam, the Netherlands (former affiliation)

