## Supplementary figures and images for "A 5-transcript signature for discriminating viral and bacterial etiology in pediatric pneumonia"

### Figure S1

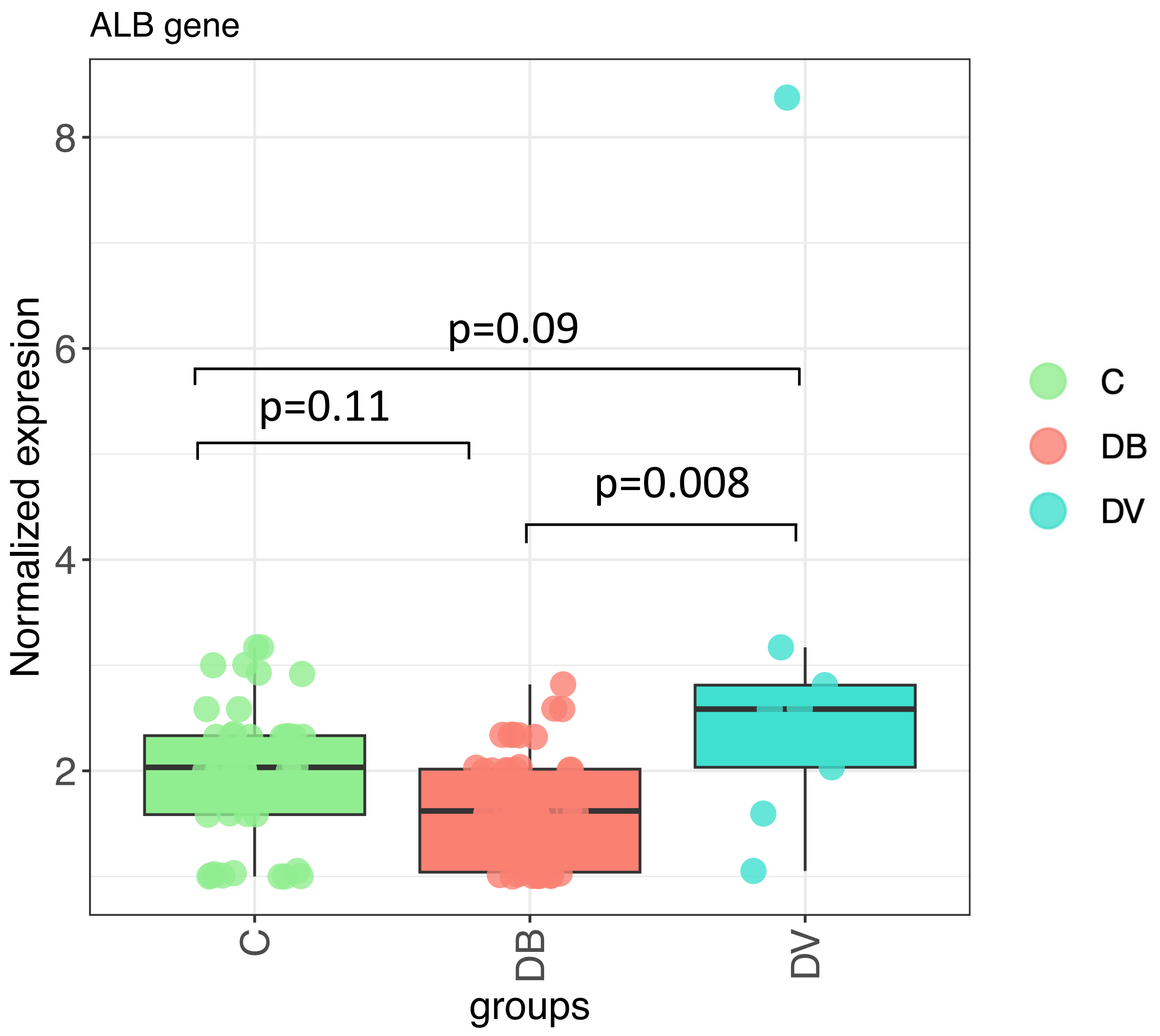

### Figure S2

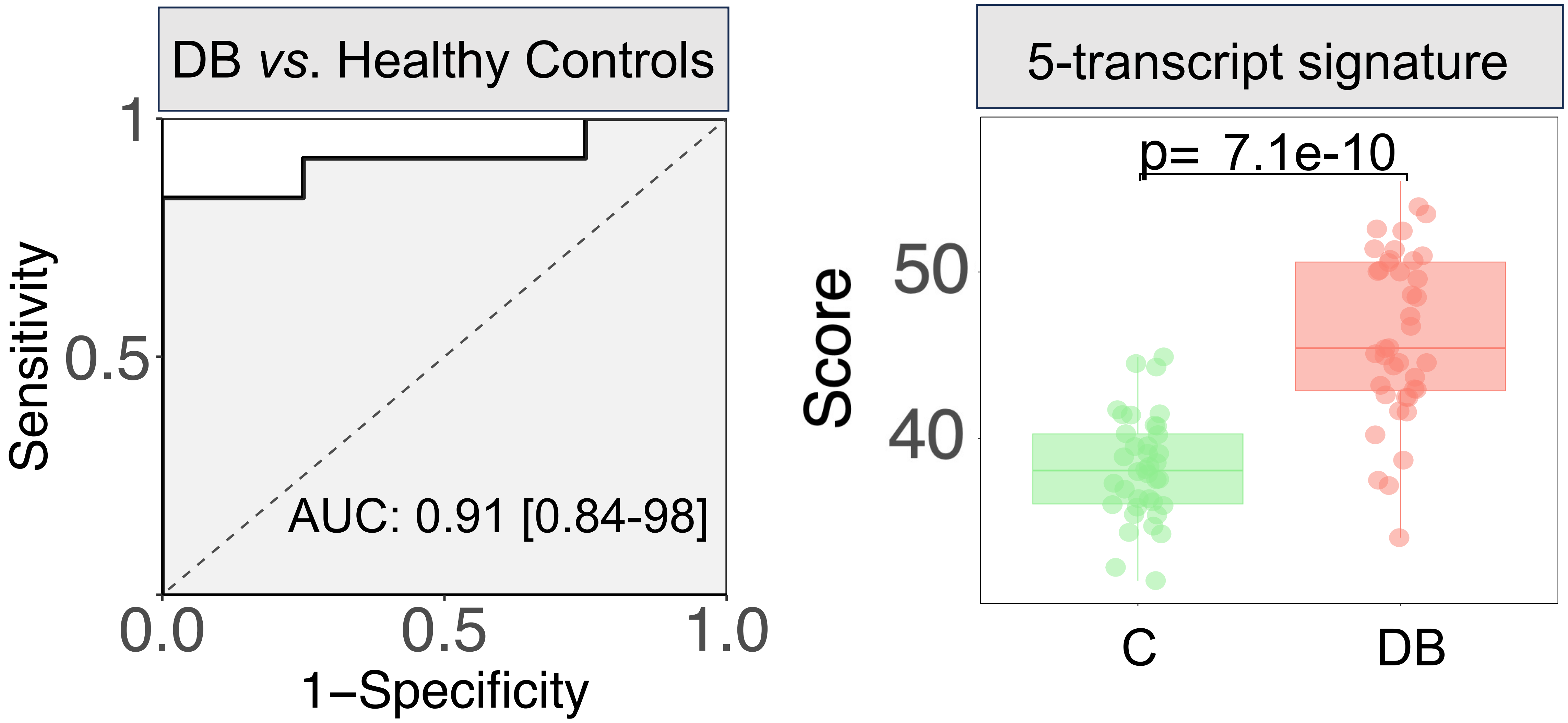

### Figure S3

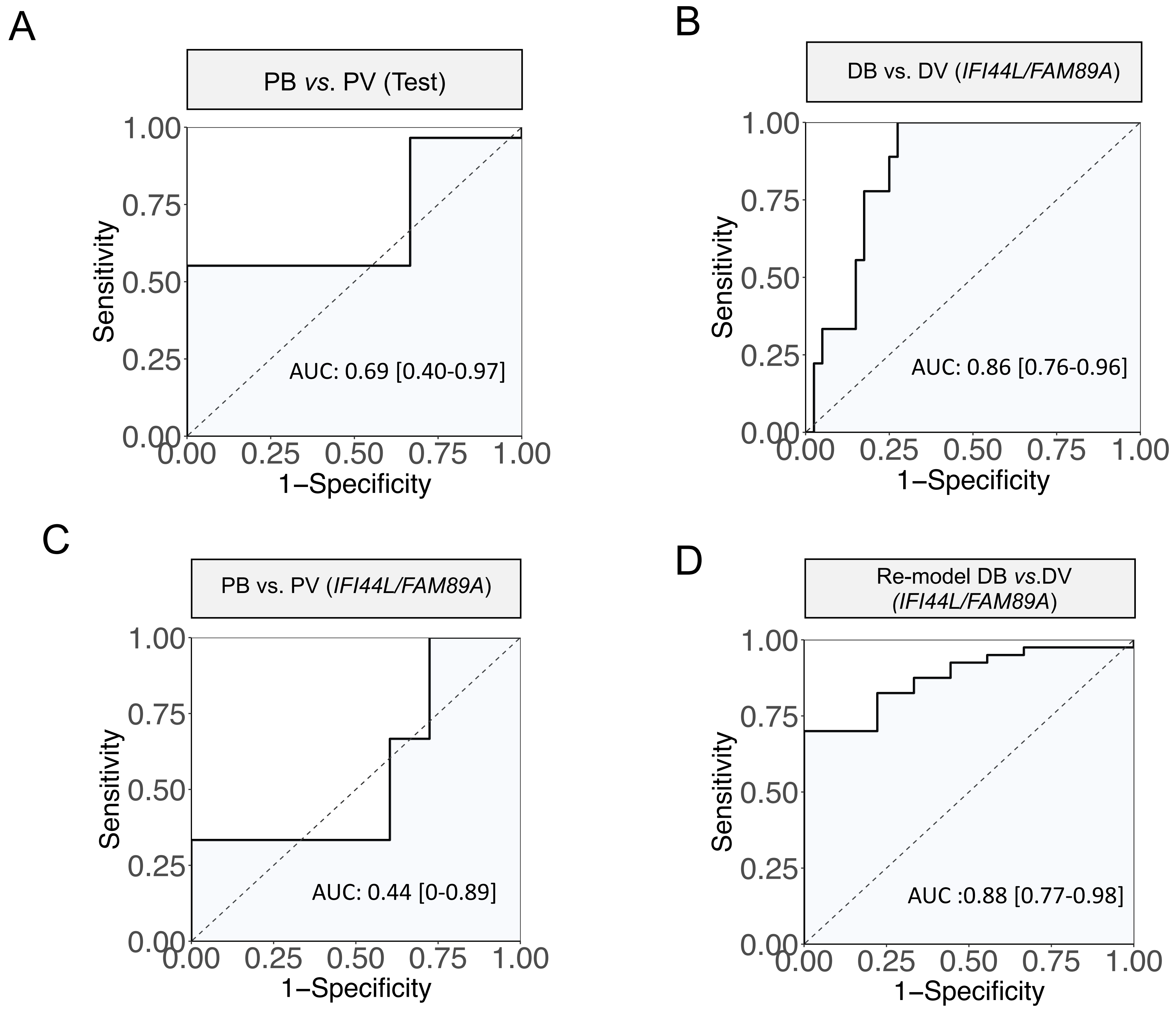

### Figure S4

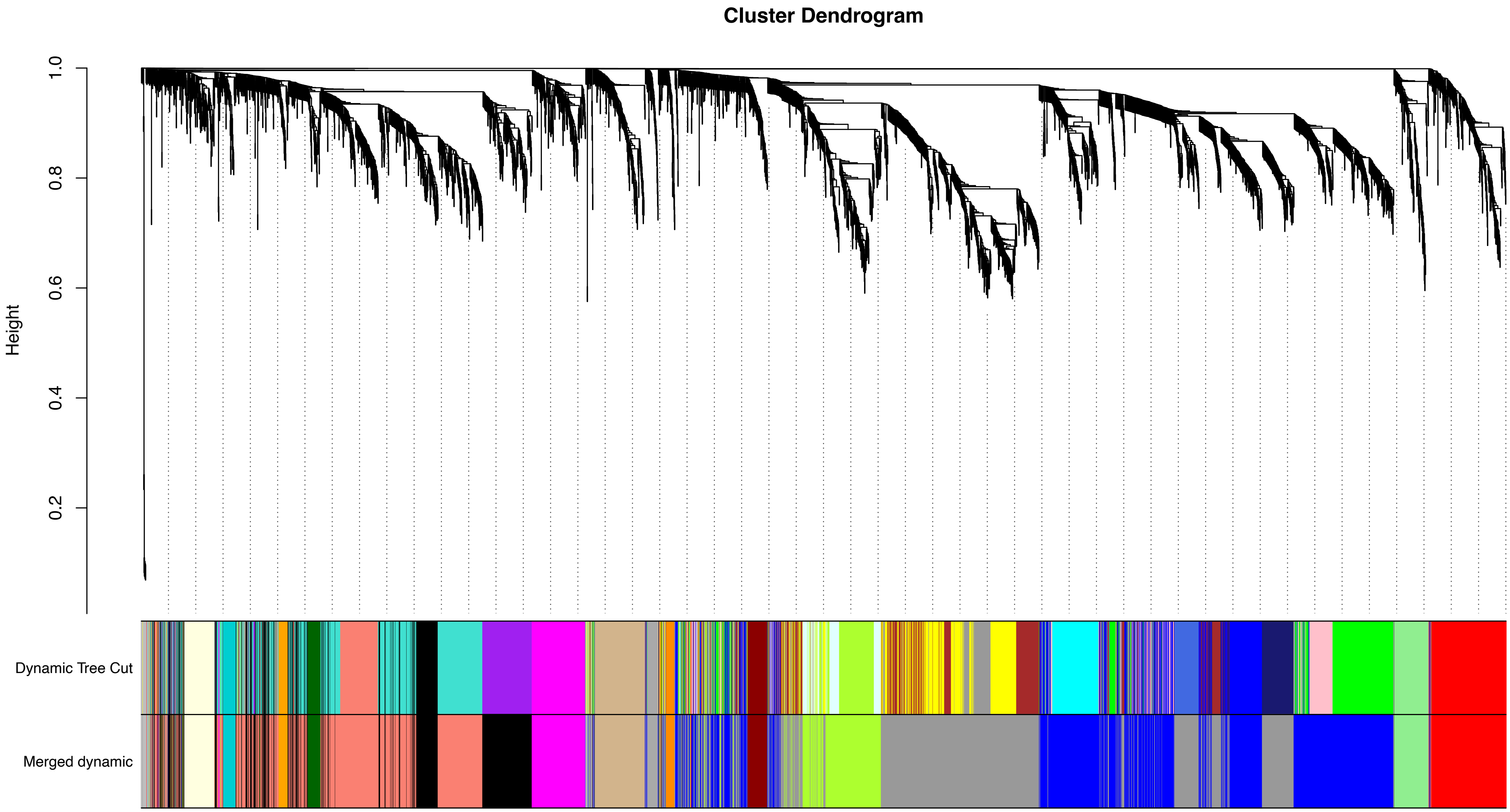

### Figure S5

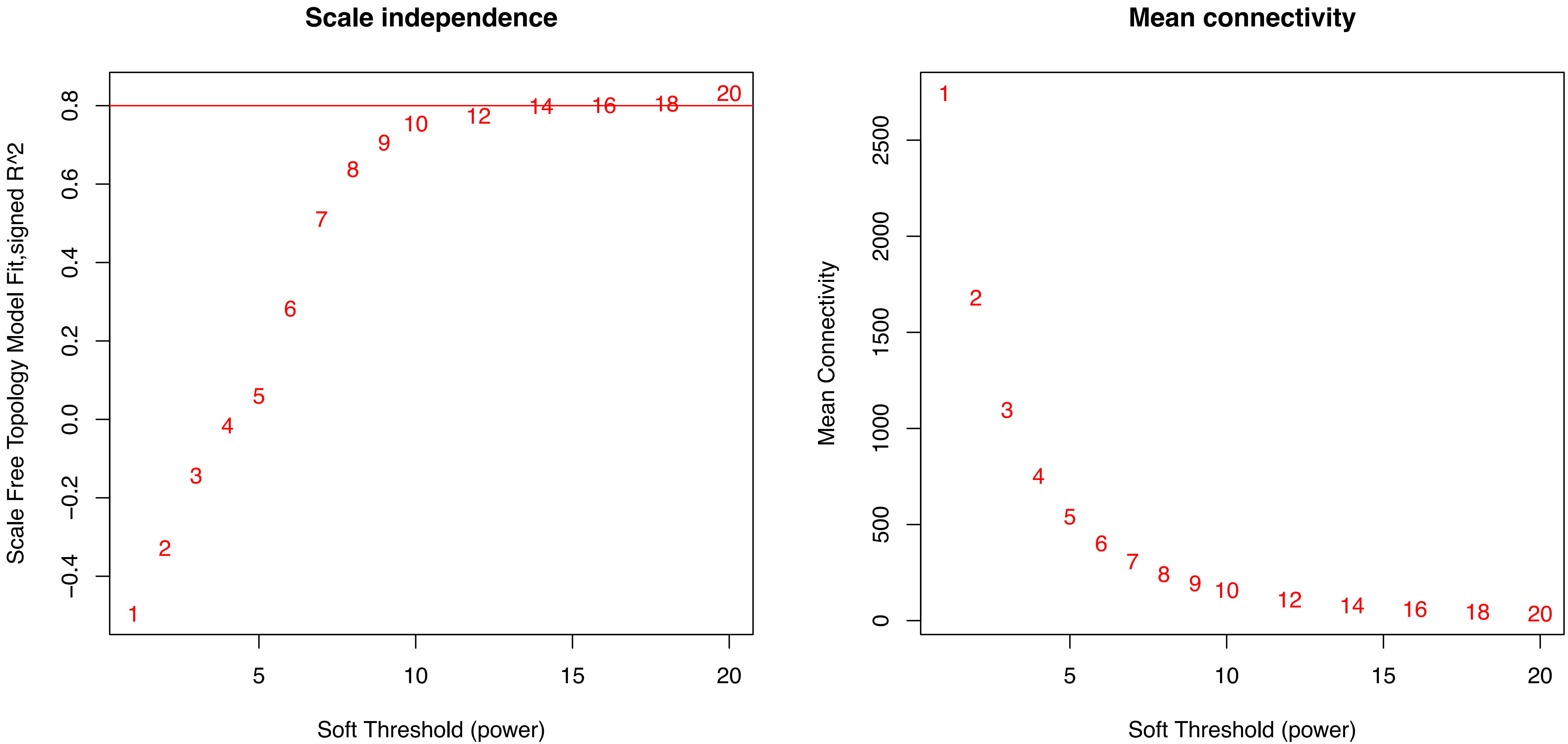
