## Supplementary material for "A 5-transcript signature for discriminating viral and bacterial etiology in pediatric pneumonia": Key Resources Table

### TABLE FOR AUTHOR TO COMPLETE

***Please do not add custom subheadings.*** *If you wish to make an entry that does not fall into one of the subheadings below, please contact your handling editor or add it under the “other*” subheading*.* ***Any subheadings not relevant to your study can be skipped.*** *(****NOTE:*** *references should be in numbered style, e.g., Smith et al.^1^)*

**Key resources table**

| REAGENT or RESOURCE | SOURCE | IDENTIFIER |
| --- | --- | --- |
| **Antibodies** | | |
| **Bacterial and virus strains** | | |
| **Biological samples** |  |  |
| Human whole blood samples | This paper | Available in Table S7 |
| **Chemicals, peptides, and recombinant proteins** | | |
| **Critical commercial assays** |  |  |
| TruSeq RNA Sample Preparation Kit | Illumina | # RS-122-2001 |
| Illumina® Ribo-Zero Gold kit | Illumina | # 20037135 |
| **Deposited data** | | |
| RNA-Seq data | This paper | GEO database: GSE261482 |
| RNA-Seq data | This paper | ArrayExpress database: E-MTAB-14564 |
| RNA-Seq data | Jackson et al.^121^ | ArrayExpress database: E-MTAB-12793 |
| RNA-Seq data | Habgood-Coote et al.^120^ | ArrayExpress database: E-MTAB-11671 |
| Microarray data | Wallihan et al. ^68^ | GEO accession number: GSE103119 |
| R code | This paper | https://github.com/Sandraviz/Pneumonia_Transcriptomic-Analysis |
| **Experimental models: Cell lines** |  |  |
| **Experimental models: Organisms/strains** |  |  |
| **Oligonucleotides** | | |
| **Recombinant DNA** | | |
| **Software and algorithms** | | |
| R software | https://www.r-project.org/ | Version 4.2.0 |
| *FastQC* | Andrews et al. ^55^ | https://github.com/s-andrews/FastQC |
| STAR | Dobin et al. ^56^ | https://github.com/alexdobin/STAR |
| FeatureCounts | Liao et al. ^58^ | https://www.bioconductor.org/packages/release/bioc/html/Rsubread.html |
| *DESeq2* | Love et al. ^59^ | https://bioconductor.org/packages/release/bioc/html/DESeq2.html |
| *RUVSeq* | Risso et al. ^60^ | https://bioconductor.org/packages/release/bioc/html/RUVSeq.html |
| *limma* | Ritchie et al. ^62^ | https://bioconductor.org/packages/release/bioc/html/limma.html |
| *Clusterprofiler* | Yu et al. ^63^ | https://bioconductor.org/packages/release/bioc/html/clusterProfiler.html |
| *glmnet* | Friedman et al. ^64^ | https://cran.r-project.org/web/packages/glmnet/index.html |
| *pROC* | Robin et al. ^65^ | https://cran.r-project.org/web/packages/pROC/index.html |
| *OptimalCutPoints* | López-Ratón et al. ^66^ | https://cran.r-project.org/web/packages/OptimalCutpoints/index.html |
| *WGCNA* | Langfelder et al. ^67^ | https://cran.r-project.org/web/packages/WGCNA/index.html |
| **Other** |  |  |
